# Integrative Genome-Wide Association Studies of COVID-19 Susceptibility and Hospitalization Reveal Risk Loci for Long COVID

**DOI:** 10.1101/2025.02.13.25322223

**Authors:** Zhongshan Cheng

## Abstract

Long COVID presents a significant public health challenge, characterized by over 200 reported symptoms spanning multiple organ systems, and genome-wide association studies (GWAS) have been hindered by the disorder’s symptom heterogeneity and the limited power of available datasets. To overcome the challenges of limited sample size or heterogeneity of long COVID cohorts, a proxy-based, hypothesis-generating strategy is conducted to prioritize candidate risk loci that may contribute to long COVID by analyzing GWAS summary statistics for COVID-19 susceptibility and hospitalization, as well as long COVID from the COVID-19 Host Genetics Initiative (HGI, Release 7), resulting in 62 candidate loci represented by independent single nucleotide polymorphisms (SNPs). These SNPs are categorized into three groups based on their association with acute-phase COVID-19: (1) severe COVID-19-specific SNPs, such as SNPs mapped to two well-known loci involved in SARS-CoV2 entry (*ACE2* [rs190509934] and *TMPRSS2* [rs12329760]), as well as variants of *DPP9* (rs7251000), *FOXP4* (rs12660421) and *HLA-DQA1* (rs17219281), exhibiting associations with severe COVID-19 but displaying weaker signals in non-hospitalized COVID-19 cases; (2) SNPs associated with both severe and mild COVID-19, including the SNP close to *ABO* (rs505922); and (3) non-hospitalization-specific SNPs, such as variants in *KCTD16* (rs62401842) and *WASF3* (rs56143829), highlighting genetic contributors specific to mild COVID-19 cases. These candidate SNPs are investigated with recently published long COVID GWAS data from HGI, revealing most of these candidate SNPs display much weaker association with long COVID. Further transcriptome-wide association studies (TWASs) for GWASs of COVID-19 hospitalization/susceptibility/long COVID across 48 GTEx tissues demonstrate that genes adjacent to these candidate SNPs (43 out of 62 SNPs) exhibits relatively weaker association signals in long COVID across heart, brain, and muscle-related tissues, particular for notable TWAS hits, *DPP9* (rs7251000), *CCR1* (rs17078348), and *THBS3* (rs41264915) that display much weaker association in long COVID compared to acute-phase COVID-19. Phenome-wide TWASs also link *HLA-DQA1* (rs17219281), *HLA-A* (rs9260038), and *HLA-C* (rs1634761), *GSDMB* (rs9916158) and *FOXP4* (rs12660421) with other phenotypes closely relevant to long COVID. These results provide new insights to long COVID by leveraging acute-phase COVID-19.

## INTRODUCTION

Long COVID remains as a critical and complex public health issue, with a diverse array of over 200 reported symptoms affecting multiple organ systems[1]. These symptoms, including fatigue, cognitive impairment, respiratory issues, and cardiovascular problems, persist far beyond the acute phase of COVID-19 infection, usually lasting longer for 3 months and posting unique challenges for researchers to identify genetic risk factors using genome-wide association studies (GWAS), mainly because of the wide variation in symptom clusters arising from different biological mechanisms of long COVID and necessity of cohorts with large sample sizes to detect genome-wide significant associations. Due to these complexity, long COVID GWAS requires not only large and diverse patient datasets but also refined approaches to phenotype classification and statistical analyses.

Among the most significant efforts to understand the genetic basis of long COVID was the large GWAS from the COVID-19 Host Genetic Initiatives (HGI) conducted by Lama et al.[2], which analyzed approximately 3,500 long COVID cases with only a single locus, *FOXP4*, on chromosome 17, passed the genome-wide significance with long COVID. Nevertheless, *FOXP4* variants were found to be strongly associated with both long COVID and severe COVID-19, in line with a shared genetic predisposition that may drive both conditions[3], and a stronger association of *FOXP4* with severe COVID-19 suggests that *FOXP4* may contribute primarily to risk factors influencing severe acute COVID-19 that later progress to long COVID[3]. Given that long COVID can also follow mild COVID-19, additional genetic markers specifically associated with mild COVID-19 that is also highly linked with long COVID likely remain undiscovered.

Another long COVID GWAS conducted by Taylor et al.[4] identified further genetic factors specific to a subtype of long COVID by investigating genetic overlap between long COVID and myalgic encephalomyelitis/chronic fatigue syndrome (ME/CFS), a condition with substantial symptom overlap, including chronic fatigue and cognitive impairments. However, this long COVID GWAS was limited to its relatively small sample size (*n* < 1,000 long COVID cases) and only reported SNPs derived by network analysis without identifying any SNPs that reached genome-wide significance, emphasizing the need for larger, more statistically robust studies.

More recently, a multi-ancestry GWAS was conducted by Chaudhary et al.[5], including participants from diverse ancestral backgrounds. Notably, several immune-related loci, such as *ABO*, *BPTF*, and *HLA-DQA1*, were identified to be linked with chronic fatigue syndrome (CFS), fibromyalgia, and depression, suggesting potential immune dysregulation in long COVID. Despite this, multi-ancestry long COVID GWAS still has limited non-European samples, leading to the lack of sufficient power to detect associations for less common symptom clusters or subtle genetic variants in non-European populations.

Recent studies on long COVID revealed that multiple risk factors, such as old age [6], female sex [7, 8], and other comorbidities [9], including both acute COVID-19 susceptibility/severity and even mild COVID-19 [6, 10], were associated with long COVID, strongly suggesting genetic factors predisposing to these conditions may also associate with long COVID. Given there were large numbers of GWASs reported to these conditions linked with long COVID, it would be feasible to integrate these GWASs to prioritize genetic factors potentially relevant to long COVID.

In fact, compared to the limited number of long COVID GWAS hits reported, large-scale GWASs from the HGI have robustly identified genetic variants associated with COVID-19 susceptibility (infection vs. population) and hospitalization (hospitalized vs. non-hospitalized or general population)[11]. Since both COVID-19 acute severity and susceptibility are known risk factors for long COVID [9], we hypothesized that genetic variants associated with these acute-phase COVID-19 may also influence the risk of developing long COVID given the strong phenotype-wide correlation between long COVID and acute-phase COVID-19[10]. To explore the rich data of acute-phase COVID-19 and search for potential long COVID risk loci, we performed an integrative analysis using COVID-19 susceptibility and hospitalization GWAS summary statistics as proxy phenotypes along with published long COVID GWAS from HGI. We first classified associated SNPs by their specificity/commonality to COVID-19 hospitalization or non-hospitalization and prioritized candidates SNPs for further evaluation of genes adjacent to them in published long COVID GWASs. Additionally, genes close to the candidate SNPs were evaluated in transcriptome-wide association studies (TWAS) of acute-phase COVID-19 and long COVID GWASs, as well as in phenome-wide association studies (PheWAS). Our objective was to prioritize a set of candidate SNPs/genes for future investigation of long COVID.

## MATERIALS AND METHODS

### Genome-wide Screening for SNPs Associated with COVID-19 Susceptibility, Hospitalization or Non-Hospitalization

An integrative workflow was developed to screening for unique/common genetic elements predisposing to COVID-19 susceptibility, hospitalization and non-hospitalization (Fig. 1). Three COVID-19 GWAS datasets with large sample sizes were downloaded from the HGI (release 7), including HGI-B1 (hospitalized [*n* = 16,512] vs. non-hospitalized COVID-19 [*n* = 71,321]), HGI-B2 (hospitalized COVID-19 [*n* = 44,986] vs. the general population [*n* = 2,356,386]), and HGI-C2 (COVID-19 [*n* = 159,840] vs. the general population [*n* = 2,782,977]). These three GWASs represented related but biologically distinct COVID-19 phenotypes. HGI-C2 primarily captures genetic susceptibility to SARS-CoV-2 infection because all infected individuals were treated as cases regardless of disease severity. In contrast, HGI-B1 isolated genetic determinants of disease severity by directly comparing hospitalized versus non-hospitalized infected individuals, thereby minimizing the contribution of infection susceptibility itself. HGI-B2 represented a composite phenotype because hospitalized COVID-19 cases were compared against the general population, comprising both uninfected individuals and infected individuals with varying disease severities. Consequently, association signals detected in HGI-B2 may reflect a combination of susceptibility to infection and progression to severe disease.

Because of this mixed phenotype structure, direct comparison among HGI-B1, HGI-B2, and HGI-C2 enables partial decomposition of genetic effects associated with susceptibility, severe disease progression, and non-hospitalized COVID-19. In particular, comparison between HGI-B1 and HGI-B2 is informative because these two GWASs share the similar hospitalized cases but differ substantially in control selection. HGI-B1 uses non-hospitalized COVID-19 individuals as controls, whereas HGI-B2 uses the general population as controls. Therefore, SNPs displaying differential effect sizes between HGI-B1 and HGI-B2 are likely influenced by genetic factors enriched in non-hospitalized COVID-19 or mild disease phenotypes.

The first two GWASs (HGI-B1 and HGI-B2) were compared for effect sizes of each SNP genome-wide using differential z-score method by considering potential sample overlap[12, 13], resulting in a new GWAS, called HGI-B1 vs. B2 (HGI-B1-vs-B2). Under this framework, SNPs displaying stronger association signals in HGI-B1 than in HGI-B2 were hypothesized to represent loci potentially enriched among non-hospitalized or mild COVID-19 phenotypes. Nevertheless, loci influenced by the presence of non-hospitalized infected individuals in the HGI-B1 control group may exhibit differential effect sizes between the two GWASs. Because HGI-B2 uses the general population as controls, this comparison may additionally capture susceptibility-related effects. Therefore, the HGI-B1-vs-B2 GWAS was interpreted as a prioritization strategy for loci associated with differential COVID-19 clinical presentation rather than a direct GWAS of mild COVID-19.

HGI also conducted long COVID GWASs with 4 slightly different case-control designs that were published by Lammi et al[14], including (1) LongCOVID-N1: long COVID strict cases (long COVID with verified SARS-CoV-2 infection) vs. strict controls (individuals with SARS-CoV-2 infection but did not have long COVID), (2) LongCOVID-N2: long COVID strict cases vs. general population (all individuals included in the cohort without long COVID), (3) LongCOVID-W1: the broad long COVID cases (long COVID after any reported SARS-CoV-2 infection) vs. the strict controls, and (4) LongCOVID-N2: the broad long COVID cases vs. general population. These long COVID GWASs were downloaded and only used in evaluation of top hits derived from the comparison of the 4 acute-phase COVID-19 GWASs.

Subsequently, by comparing these acute-phase COVID-19 GWASs, SNPs were separated into three categories, with the first categories labeled as “Severe COVID-19-Specific SNPs,” which includes SNPs associated with both SARS-CoV-2 susceptibility and COVID-19 hospitalization but tends to be more prevalent within hospitalized than in non-hospitalized COVID-19, the second SNP category named as “SNPs Associated with Both Severe and Mild COVID-19,” which influences both hospitalized and non-hospitalized COVID-19, and the last SNP category defined as “Mild COVID-19-Specific SNPs,” which tends to be specifically associated with non-hospitalized COVID-19.

For the 1^st^ SNP category, “Severe COVID-19-Specific SNPs,” the following selection criteria were applied, including (a) *P* ≤ 0.05 in HGI-C2, capturing SNPs linked to COVID-19 susceptibility; (b) *P* > 0.05 in the differential GWAS of HGI-B1 vs. HGI-B2, keeping SNPs without differential effect sizes between these comparisons; this filter is set based on the consideration that current sample sizes of these COVID-19 GWASs didn’t have enough power to detect differential effect sizes for these severe COVID-19 specific SNPs displaying stronger association between hospitalized (severe cases) and non-hospitalized (mild cases) COVID-19 than that of association between hospitalized COVID-19 and general population; (c) *P* ≤ 0.05 in HGI-B2, identifying SNPs distinguishing hospitalized COVID-19 individuals from the general population; (d) *P* ≤ 0.05 in HGI-B1, identifying SNPs that predispose individuals to COVID-19 hospitalization rather than non-hospitalized COVID-19; (e) all SNPs required to display consistent effect size directions across HGI-C2 and HGI-B2. Only independent SNPs passed the *P <* 1×10^-7^ in any of the three GWASs (HGI-B1, HGI-B2, or HGI-C2) were selected as candidate SNPs.

In terms of the 2^nd^ SNP category, “SNPs Associated with Both Severe and Mild COVID-19,” representing SNPs not significant between hospitalized and non-hospitalized cases but exhibiting associations with SARS-CoV-2 susceptibility and COVID-19 hospitalization when compared to the general population. The selection criteria for this group were performed as follows: (a) *P* ≤ 0.05 in HGI-C2, identifying COVID-19 susceptibility-related SNPs; (b) *P* ≤ 0.05 in the differential GWAS of HGI-B1 vs. HGI-B2, capturing SNPs with distinct effects in the two GWASs; (c) *P* ≤ 0.05 in HGI-B2, identifying SNPs associated with COVID-19 hospitalization; (d) *P* > 0.05 in HGI-B1, capturing SNPs with non-significant difference in effect sizes between severe and mild COVID-19; this is an important filter to confirm that the tested SNP demonstrates significant association when comparing hospitalized COVID-19 with general population but displays non-significant distribution between hospitalized and non-hospitalized COVID-19 mainly because of the nature of the SNP associated with both severe and mild COVID-19. All prioritized SNPs need to fulfill consistent effect size directions across HGI-C2 and HGI-B2. Similarly, only independent SNPs passing the *P* < 1×10^-7^ in any of these three GWASs (HGI-B1, HGI-B2, or HGI-C2) were selected as candidate SNPs.

The last SNP category was designated as “Mild COVID-19-Specific SNPs.” The rationale underlying this category is that most existing HGI GWASs were optimized to detect susceptibility or hospitalization-associated loci, potentially reducing sensitivity for loci enriched among non-hospitalized COVID-19 individuals. We therefore aimed to prioritize SNPs showing differential association patterns between HGI-B1 and HGI-B2 that may reflect genetic effects more prominent in non-hospitalized or milder COVID-19 phenotypes. To screening for mild-COVID-19 biased SNPs, we applied the following criteria to prioritize SNPs: (a) *P* ≤ 0.05 in the differential GWAS between HGI-B1 and HGI-B2, retaining SNPs with differential association signals between these datasets; this is the most important filter to obtain SNPs showing different effect size between the two GWASs mainly due to the utilization of two slightly different controls, i.e., non-hospitalized COVID-19 used as controls for HGI-B1 and general population treated as controls for HGI-B2; (b) *P* > 0.05 in the GWAS comparing hospitalized cases with the general population (HGI-B2), which excludes SNPs associated with COVID-19 hospitalization; this filter ensures selected SNPs associated with mild COVID-19 but not severe COVID represented by the hospitalization status; (c) *P* ≤ 0.05 in the GWAS comparing hospitalized cases with non-hospitalized cases (HGI-B1), which captures SNPs significantly differing these two groups, ensuring SNPs specifically associated with mild COVID-19. To further refine the selection, independent SNPs with minor allele frequency (MAF) > 0.01 in the largest cohort HGI-C2 (without filtering by significance) and met a suggestive significance threshold (*P* < 1×10^-5^) in any of HGI-B1 and HGI-B1-vs-B2 were prioritized. These SNPs were also ensured that no other genome-wide significant SNPs close to it within a 500-kb window in either HGI-B2 or HGI-C2. The use of a relaxed significance threshold and stricter MAF filter aimed to account for the higher heterogeneity among non-hospitalized cases in HGI-B1 and the inherent bias of HGI GWASs toward identifying SNPs associated with hospitalization, due to the relative small size of non-hospitalized COVID-19. This approach enables the identification of SNPs that might specifically associate with non-hospitalized COVID-19, although COVID-19 susceptibility specific SNPs might be included, which can be easily identified later based on these SNPs’ association signals in HGI-C2.

Finally, all these candidate SNPs obtained using the above criteria were subjected to detailed evaluation by local Manhattan plots among acute-phase COVID-19 GWASs of susceptibility and hospitalization from HGI, sex-stratification COVID-19 hospitalization GWASs from UK Biobank, and long COVID GWASs from HGI (Fig. 1).

### Comparing Two GWASs Using Differential Z-Score Method

To assess differences in effect sizes of SNPs between two GWASs, such as one GWAS comparing hospitalized cases with non-hospitalized cases (HGI-B1) and the other GWAS comparing hospitalized cases with the general population (HGI-B2), or two sex-stratified COVID-19 hospitalization GWASs for males and females, differential z-score method[12] was applied in the current study. This approach quantifies the disparity in effect sizes (β) of each SNP between two GWASs by calculating a differential z-score, which incorporates adjustments for potential sample overlap between studies. The corresponding p-value was derived from the normalized differential z-scores. The differential z-score is defined as:

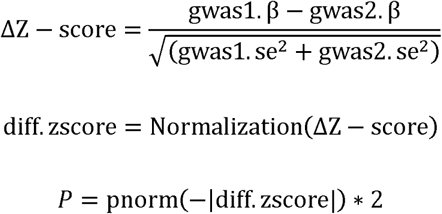

Here, gwas1.β and gwas2.β represent the effect sizes of a given SNP from the two GWASs, while gwas1.se and gwas2.se denote the corresponding standard errors. To account for genome-wide correlation in effect sizes caused by overlapping samples, differential z-scores were normalized using the SAS procedure “proc stdize” with the “std” method. Subsequently, differential p-values were computed based on the normalized differential z-scores. The cumulative distribution function of the standard normal distribution (“pnorm”) was used to calculate p-values from the normalized z-scores.

### Sex-Biased Associations with COVID-19 Hospitalization among Top Candidate SNPs

Epidemiological evidence indicates that women are more likely to experience long COVID but less likely to develop severe COVID-19[7, 8], potentially due to stronger immune responses to SARS-CoV-2 infection in women[8]. To investigate whether these candidate SNPs exhibit sex-biased associations with COVID-19 hospitalization/non-hospitalization, we analyzed publicly available sex-stratified GWAS summary statistics from the GRASP COVID-19 database[15] generated based on UK Biobank samples. The selected sex-stratified COVID-19 hospitalization GWASs (cases=1,343 and controls=262,886 for females; cases=1,917 and controls=221,174 for males) includes mixed ancestry populations which are similar to the four HGI COVID-19 GWAS datasets used in our study. We used the aforementioned differential z-score method to compare male and female COVID-19 hospitalization GWAS results, and SNPs with differential effect sizes between sexes for COVID-19 hospitalization were identified. Genomic inflation factors for both sex-stratified GWASs and the sex-biased differential GWAS were also evaluated to ensure no inflations observed among them.

### Evaluating the Associations of Top Candidate SNPs and Its Adjacent SNPs with Long COVID

Four HGI long COVID GWASs, including LongCOVID-N1, LongCOVID-N2, LongCOVID-W1, and LongCOVID-W2, with slightly different case and control designs were used to evaluate the association signals of these candidate SNPs. Due to the nature of phenotypic differences between acute-phase COVID-19 and long COVID, adjacent SNPs, located within a 1Mb-genomic window where each top candidate SNP residing in, were screened across these long COVID GWASs, and the one with the smallest long COVID association signals passed the significance threshold of *P <* 0.001 were kept for further analyses, such as manual evaluation of local Manhattan plots and forest plots for these candidate variants. These loci with nominally significant top SNPs or its adjacent SNPs passed the arbitrary prioritization threshold of *P <* 0.001 in any of these long COVID GWASs were treated as potential candidate loci for long COVID.

### Evaluating Previously Reported Long COVID SNPs in Current Acute-Phase COVID-19 GWASs

To assess whether these prioritized candidate SNPs identified in our study overlap with or are proximal to those reported in two independent long COVID GWASs by Taylor et al.[4] and Chaudhary et al.[5], a set of 83 SNPs derived from SNP network analyses and 3 SNPs identified via traditional GWAS methods specifically from the two studies were created and analyzed for their association signals among the three acute-phase GWASs from HGI.

### Transcriptome-Wide Association Analysis Using S-MetaXcan

TWAS was conducted using S-MetaXcan[16], which infers gene expression association with a phenotype based on its corresponding GWAS summary statistics and GTEx gene expression datasets across 48 tissues. S-MetaXcan is able to integrates GWAS summary statistics with expression quantitative trait loci (eQTL) reference models derived from GTEx tissues to estimate the association between genetically regulated gene expression and a phenotype of interest, which leverages pre-trained prediction models generated from GTEx data, and the expression level of each gene within a specific tissue is modeled as a weighted combination of multiple cis-eQTL SNPs located near that gene. The weights assigned to each SNP are estimated using elastic net regression based on GTEx genotype-expression reference datasets, allowing multiple eQTLs with independent or correlated effects to jointly contribute to the prediction of gene expression. Base on provided GWAS summary statistics, S-MetaXcan then aggregates SNP-level association signals according to these tissue-specific eQTL weights while accounting for linkage disequilibrium (LD) among SNPs through an external LD reference panel, such as the European population from the 1000 Genome Project. This framework estimates whether the genetically predicted expression level of a gene is significantly associated with the trait investigated by the GWAS. Because prediction models are generated independently across multiple GTEx tissues, S-MetaXcan enables tissue-specific evaluation of gene-trait associations and facilitates identification of genes whose genetically regulated expression may contribute to disease risk in biologically relevant tissues.

Four COVID-19 GWAS datasets from the HGI were analyzed in TWAS, including HGI-B1, HGI-B2, HGI-C2, and HGI-B1-vs-B2, along with four long COVID GWAS datasets with subtly distinct case-control definitions from HGI[14] and an additional set of 222 GWASs from the UK Biobank (UKB) (all sample sizes of cases >5000) shared by Benneal’s lab (https://www.nealelab.is/uk-biobank). Reference transcriptomic data from 48 tissues in GTEx v7[17] were utilized for gene expression imputation in the analyses.

At the beginning, we first evaluated the TWASs of 4 acute-phase COVID-19 using an arbitrary prioritization threshold, i.e., *P* = 3.5×10⁻[. This arbitrary threshold was used to prioritize adjacent genes for future investigation of long COVIDs, which was calculated based on the total number of GTEx tissues (n=48), total number of adjacent genes (n=753), total number of HGI-GWASs (n=4; HGI-B1, HGI-B2, HGI-C2, and HGI-B1-vs-B2), using the strict Bonferroni correction method with an arbitrary relaxed scale factor (c=10) considering the potential correlation of association among these 48 GTEx tissues, i.e., 10*0.05/(753*48*4).

In order to incorporate the long COVID TWASs into the acute-phase COVID-19 TWASs, TWAS results of COVID GWAS datasets (*n* = 8, collectively referred to as COVID TWASs, covering 4 long COVID TWASs as well as 4 acute-phase COVID-19 TWASs) were evaluated for genes proximal to prioritized SNPs (±500kb to each SNP termed as tag SNP). Any genes showing nominal significance (*P <* 0.05) in any COVID TWASs across the 48 GTEx tissues were included as supplementary data for further investigation. This is also an arbitrary threshold to prioritize adjacent genes, as most of them demonstrate much weaker TWAS association signals in long COVID compared to acute-phase COVID-19.

Also, to expand these findings, phenome-wide association analyses were conducted for these prioritized genes. Associations were examined across a TWAS database aggregating 190 previously published TWAS results covering a broad set of phenotypes[16] and 222 UKB TWASs created locally with the same settings as that of the 190 published TWASs, as well as 8 COVID related TWASs generated in this study, yielding a total of 420 TWAS datasets (see Table S7).

### Statistical Analysis Software and Availability of Codes and Data

Unless otherwise specified, all analyses were performed using SAS OnDemand for Academics (freely available at https://www.sas.com/en_us/software/on-demand-for-academics.html). Local Manhattan plots for the prioritized candidate SNPs are publicly accessible at the GitHub link (https://github.com/chengzhongshan/COVID19_GWAS_Analyzer/tree/main/LongCOVID_data_and_scripts/LongCOVID_SNP_LocalManhattanPlots). SAS scripts involved in post-GWAS analyses for these acute-phase COVID-19 and long COVID GWASs are deposited into the same GitHub folder. Acute-phase COVID-19 and long COVID GWASs used in our study are available via Google Driver (THREEGWASs4HGICOVID19.gz: https://drive.google.com/file/d/1nZZsxbXJROjGmBGqq84WbCtPLlZWDT3C/view?usP; CombineLongCOVIDGWAS.gz: https://drive.google.com/file/d/1opd3BSC5JAHoPvDZPmqPB11Ox8YTrG-X/view?usP). A readme document explaining details of the two compressed datasets is provided at (https://docs.google.com/document/d/1EVA-gUZyGeiyb4GPDuliUyT044WBya_I).

## RESULTS

### Comparison of Three Acute-Phase COVID-19 GWASs from HGI

To search for candidate genetic SNPs commonly or specifically associated with COVID-19 hospitalization and non-hospitalization, an integrative workflow was developed by leveraging publicly available COVID-19 GWAS (Fig. 1). Initially, acute COVID-19 phenotypes, such as SARS-CoV-2 susceptibility, COVID-19 hospitalization, and non-hospitalization, were used to prioritize genetic variants potentially and specifically linked to COVID-19 hospitalization or non-hospitalization, as well as associated with both of the two COVID-19 phenotypes. The analysis utilized GWAS data from the HGI release 7, including GWASs of hospitalized vs. non-hospitalized cases (HGI-B1), hospitalized cases vs. the general population (HGI-B2), and SARS-CoV-2 infection vs. the general population (HGI-C2). These GWASs comprise large sample sizes (16,512, 44,986, and 159,840 COVID-19 cases for HGI-B1, HGI-B2, and HGI-C2, respectively; Fig. 1) and exhibit minimal genomic inflation (all λs close to 1), as shown in supplementary Fig. S1, and two loci, including *SLC6A20* and *ABO*, displayed notable genome-wide significance, which were well-established loci associated with COVID-19 susceptibility and severity[18].

### Candidate SNPs Specifically Associated with Hospitalized COVID-19 or Commonly Associated with Both Hospitalized and Non-hospitalized COVID-19

Base on the filtering strategies illustrated in the workflow shown in Fig. 1 (see detail in Method section), two SNP groups were prioritized first, including “Severe COVID-19-Specific SNPs” (*n* = 27) and “SNPs Associated with Both Severe and Mild COVID-19” (*n* = 15), resulting in 42 independent SNPs (Table 1 and Fig. 2). It is necessary to emphasize that these two SNP categories were designated to capture SNPs either linked predominantly to SARS-CoV-2 susceptibility and COVID-19 hospitalization coupled with higher prevalence in hospitalized cases than in non-hospitalized cases, or associated with both severe and mild COVID-19 that demonstrate consistent associations with SARS-CoV-2 susceptibility and hospitalization along with no frequency difference between hospitalized and non-hospitalized cases. In addition, it is also necessary to notice that HGI-B2 captures both susceptibility and severity components, and SNP categorization was based on comparative signal patterns across all three GWASs rather than interpretation from any single dataset alone. Among these 42 independent SNPs included in the two SNP categories, all SNPs passed the conventional genome-wide significance threshold (*P <* 5×10^-8^) in at least one HGI COVID-19 GWAS, with the exception of two SNPs, rs150345524 (*SF3B1*, *P =* 5.6×10^-8^ in HGI-B2) and rs11766643 (*SLC22A5*, *P* = 7.5×10^-8^ in HGI-C2), that are suggestively genome-wide significant and were manually selected after viewing association signals around them in local Manhattan plots and also based on its potential involvement in COVID-19.

**Table 1.** Candidate SNPs associated with either COVID-19 hospitalization or both COVID-19 hospitalization and non-hospitalization.

| Variant information |  |  |  |  |  | HGI-B1 |  |  | HGI-B2 |  |  | HGI-C2 |  |  | HGI-B1-vs-B2 | COVID-19 |
| --- | --- | --- | --- | --- | --- | --- | --- | --- | --- | --- | --- | --- | --- | --- | --- | --- |
| chr | pos | rsid | allele | AF | gene | beta | se | p | beta | se | p | beta | se | p | DiffPval | SNP groups |
| 1 | 47275770 | rs10890422 | C | 0.402 | *CYP4B1* | -0.05 | 0.02 | 3.28E-03 | -0.05 | 0.01 | 4.43E-08 | -0.02 | 0.004 | 1.10E-04 | 6.63E-01 | Hosp |
| 1 | 65412830 | rs11208552 | T | 0.647 | *JAK1* | -0.01 | 0.02 | 6.95E-01 | -0.05 | 0.01 | 2.19E-09 | -0.02 | 0.004 | 4.21E-04 | 5.14E-03 | Both |
| 1 | 77949123 | rs7515509 | A | 0.366 | *AK5* | 0.08 | 0.02 | 5.71E-06 | 0.06 | 0.01 | 2.94E-12 | 0.02 | 0.004 | 2.16E-05 | 1.82E-01 | Hosp |
| 1 | 155167786 | rs41264915 | G | 0.103 | *THBS3* | -0.10 | 0.03 | 3.59E-04 | -0.13 | 0.01 | 9.33E-23 | -0.02 | 0.007 | 1.13E-03 | 2.64E-01 | Hosp |
| 2 | 26833309 | rs1275969 | T | 0.560 | *CIB4* | 0.04 | 0.02 | 2.37E-02 | 0.05 | 0.01 | 2.03E-08 | 0.01 | 0.004 | 9.18E-03 | 8.15E-01 | Hosp |
| 2 | 60707588 | rs1123573 | G | 0.367 | *BCL11A* | -0.03 | 0.02 | 1.86E-01 | -0.07 | 0.01 | 1.32E-14 | -0.03 | 0.004 | 4.62E-09 | 1.84E-02 | Both |
| 2 | 198245297 | rs150345524 | A | 0.413 | *SF3B1* | 0.05 | 0.02 | 1.69E-02 | 0.05 | 0.01 | 5.56E-08 | 0.01 | 0.004 | 4.66E-02 | 9.52E-01 | Hosp |
| 3 | 45847241 | rs17078348 | G | 0.101 | *SLC6A20* | 0.29 | 0.03 | 1.15E-19 | 0.34 | 0.01 | 2.53E-130 | 0.08 | 0.007 | 2.01E-36 | 6.34E-02 | Hosp |
| 3 | 101525625 | rs2290859 | T | 0.341 | *NXPE3* | -0.03 | 0.02 | 1.55E-01 | -0.07 | 0.01 | 2.34E-13 | -0.05 | 0.004 | 4.43E-31 | 2.02E-02 | Both |
| 4 | 25435159 | rs7684660 | C | 0.829 | *ANAPC4* | -0.06 | 0.03 | 1.98E-02 | -0.08 | 0.01 | 8.71E-12 | -0.02 | 0.005 | 2.86E-05 | 4.57E-01 | Hosp |
| 5 | 131776967 | rs10066378 | C | 0.131 | *SLC22A5* | 0.06 | 0.02 | 1.80E-02 | 0.07 | 0.01 | 1.63E-10 | 0.02 | 0.006 | 1.00E-03 | 4.94E-01 | Hosp |
| 6 | 29907544 | rs9260038 | G | 0.622 | *HLA-A* | -0.04 | 0.02 | 2.72E-02 | -0.06 | 0.01 | 1.94E-10 | -0.01 | 0.005 | 1.30E-02 | 3.40E-01 | Hosp |
| 6 | 31274027 | rs1634761 | T | 0.500 | *HLA-C* | -0.04 | 0.02 | 1.81E-02 | -0.06 | 0.01 | 4.50E-12 | -0.02 | 0.004 | 9.99E-07 | 2.95E-01 | Hosp |
| 6 | 32675645 | rs17219281 | A | 0.072 | *HLA-DQA1* | -0.10 | 0.04 | 1.01E-02 | -0.12 | 0.02 | 5.53E-11 | -0.03 | 0.008 | 1.81E-03 | 5.30E-01 | Hosp |
| 6 | 41488378 | rs12660421 | A | 0.041 | *FOXP4* | 0.26 | 0.04 | 2.88E-10 | 0.24 | 0.02 | 2.82E-31 | 0.08 | 0.011 | 7.36E-15 | 6.55E-01 | Hosp |
| 7 | 99630342 | rs2897075 | T | 0.364 | *ZKSCAN1* | 0.01 | 0.02 | 7.21E-01 | 0.05 | 0.01 | 8.92E-09 | 0.02 | 0.004 | 6.17E-06 | 7.17E-03 | Both |
| 7 | 100220850 | rs11766643 | G | 0.204 | *TFR2* | -0.05 | 0.02 | 2.45E-02 | -0.04 | 0.01 | 4.71E-04 | -0.03 | 0.005 | 7.46E-08 | 4.92E-01 | Hosp |
| 8 | 61537523 | rs1120591 | C | 0.393 | *RAB2A* | 0.02 | 0.02 | 2.75E-01 | 0.05 | 0.01 | 8.64E-10 | 0.02 | 0.004 | 2.48E-05 | 4.32E-02 | Both |
| 9 | 15795833 | rs79611697 | T | 0.068 | *CCDC171* | 0.08 | 0.04 | 3.75E-02 | 0.10 | 0.02 | 3.57E-08 | 0.03 | 0.009 | 3.26E-03 | 4.72E-01 | Hosp |
| 9 | 21172825 | rs149533170 | A | 0.009 | *IFNA21* | 0.06 | 0.11 | 6.03E-01 | 0.28 | 0.05 | 1.38E-08 | 0.08 | 0.025 | 7.64E-04 | 2.21E-02 | Both |
| 9 | 136149229 | rs505922 | T | 0.348 | *ABO* | 0.01 | 0.02 | 6.16E-01 | 0.09 | 0.01 | 4.14E-24 | 0.08 | 0.004 | 7.93E-90 | 2.33E-06 | Both |
| 10 | 81723695 | rs61860402 | T | 0.112 | *SFTPD* | 0.08 | 0.03 | 2.65E-03 | 0.08 | 0.01 | 1.38E-10 | 0.03 | 0.006 | 2.76E-08 | 9.61E-01 | Hosp |
| 11 | 1241221 | rs35705950 | T | 0.100 | *MUC5B* | -0.09 | 0.03 | 3.84E-03 | -0.10 | 0.01 | 3.41E-12 | -0.02 | 0.007 | 3.31E-03 | 6.00E-01 | Hosp |
| 11 | 34529831 | rs2924480 | C | 0.399 | *ELF5* | -0.03 | 0.02 | 1.67E-01 | -0.08 | 0.01 | 1.63E-16 | -0.02 | 0.004 | 1.59E-06 | 1.31E-02 | Both |
| 12 | 113374748 | rs10774679 | T | 0.653 | *OAS3* | 0.04 | 0.02 | 2.31E-02 | 0.07 | 0.01 | 3.25E-16 | 0.03 | 0.004 | 4.75E-10 | 7.42E-02 | Hosp |
| 12 | 133141973 | rs5023077 | C | 0.513 | *FBRSL1* | -0.05 | 0.02 | 1.03E-02 | -0.07 | 0.01 | 2.84E-15 | -0.02 | 0.004 | 3.14E-07 | 1.79E-01 | Hosp |
| 13 | 113535741 | rs12585036 | T | 0.209 | *ATP11A* | 0.07 | 0.02 | 4.58E-04 | 0.10 | 0.01 | 3.15E-21 | 0.02 | 0.005 | 2.85E-06 | 2.22E-01 | Hosp |
| 14 | 29497258 | rs150497803 | T | 0.043 | *FOXG1* | 0.00 | 0.05 | 9.79E-01 | -0.09 | 0.02 | 6.68E-05 | -0.07 | 0.011 | 2.79E-10 | 3.11E-02 | Both |
| 16 | 89262657 | rs117169628 | A | 0.135 | *SLC22A31* | 0.09 | 0.03 | 4.68E-04 | 0.10 | 0.01 | 1.37E-14 | 0.03 | 0.006 | 2.78E-05 | 7.97E-01 | Hosp |
| 17 | 38182229 | rs9916158 | T | 0.365 | *GSDMA/B* | -0.01 | 0.02 | 4.85E-01 | 0.03 | 0.01 | 2.05E-04 | 0.02 | 0.004 | 1.11E-08 | 5.88E-03 | Both |
| 17 | 44057595 | rs3785884 | A | 0.187 | *MAPT* | -0.05 | 0.02 | 2.22E-02 | -0.09 | 0.01 | 3.84E-16 | -0.02 | 0.005 | 3.51E-04 | 6.69E-02 | Hosp |
| 17 | 44787312 | rs1378358 | T | 0.167 | *NSF* | -0.04 | 0.02 | 9.81E-02 | -0.08 | 0.01 | 1.18E-13 | -0.02 | 0.005 | 2.58E-03 | 2.75E-02 | Both |
| 17 | 47950329 | rs75432325 | C | 0.042 | *TAC4* | 0.08 | 0.04 | 5.82E-02 | 0.18 | 0.02 | 2.24E-19 | 0.05 | 0.010 | 7.00E-07 | 2.04E-02 | Both |
| 19 | 4714468 | rs7251000 | T | 0.432 | *DPP9* | -0.07 | 0.02 | 3.98E-04 | -0.08 | 0.01 | 1.17E-19 | -0.03 | 0.004 | 1.18E-11 | 4.61E-01 | Hosp |
| 19 | 9009161 | rs12976386 | T | 0.552 | *MUC16* | 0.04 | 0.03 | 1.14E-01 | -0.03 | 0.01 | 9.37E-03 | -0.03 | 0.005 | 3.75E-08 | 2.43E-03 | Both |
| 19 | 10463118 | rs34536443 | C | 0.043 | *TYK2* | 0.05 | 0.06 | 3.58E-01 | 0.23 | 0.02 | 1.74E-21 | 0.05 | 0.012 | 1.29E-05 | 4.66E-04 | Both |
| 19 | 49206417 | rs492602 | G | 0.460 | *FUT2* | -0.02 | 0.02 | 1.69E-01 | -0.06 | 0.01 | 1.71E-11 | -0.04 | 0.004 | 3.44E-18 | 4.00E-02 | Both |
| 19 | 50882619 | rs1405655 | C | 0.332 | *NR1H2* | 0.07 | 0.02 | 6.03E-05 | 0.08 | 0.01 | 5.92E-19 | 0.03 | 0.004 | 1.20E-09 | 7.95E-01 | Hosp |
| 21 | 34621948 | rs2834164 | C | 0.480 | *IFNAR2* | -0.10 | 0.02 | 1.51E-08 | -0.11 | 0.01 | 4.14E-41 | -0.03 | 0.004 | 6.54E-14 | 4.83E-01 | Hosp |
| 21 | 35353264 | rs76608815 | T | 0.090 | *SLC5A3* | 0.11 | 0.03 | 9.64E-05 | 0.13 | 0.01 | 6.60E-21 | 0.04 | 0.007 | 1.49E-07 | 4.83E-01 | Hosp |
| 21 | 42852497 | rs12329760 | T | 0.258 | *TMPRSS2* | -0.07 | 0.02 | 8.60E-04 | -0.06 | 0.01 | 2.16E-08 | -0.01 | 0.005 | 5.32E-03 | 4.55E-01 | Hosp |
| 23 | 15620340 | rs190509934 | C | 0.003 | *ACE2* | -0.46 | 0.20 | 2.12E-02 | -0.44 | 0.10 | 1.08E-05 | -0.36 | 0.041 | 1.70E-18 | 9.38E-01 | Hosp |

Note: HGI: the COVID-19 host genetics initiative; chr: chromosome; pos: hg38 genomic position; allele: alternative allele; AF: allele frequency of alternative allele; diffPval: statistical significance *P* for differential effect size of each common SNP by comparing the two HGI GWASs, including HGI-B1 and HGI-B2; Hosp: SNPs tend to be more specifically emerged in hospitalized COVID-19 cases; Both: SNPs tend to be emerged in both hospitalized and non-hospitalized COVID-19 cases. Association *P*s passed the suggestive genome-wide association significance threshold of $P < 1 \times 10^{-7}$ are included along with differential effect size significance (DiffPval). Note: HGI-C2 (SASR-CoV2 susceptibility vs. general population), HGI-B1 (hospitalized vs. non-hospitalized COVID-19), and HGI-B2 (hospitalized vs. general population).

To be biologically intuitive, each of the above 42 COVID-19-associated SNPs was tagged with its corresponding nearest gene to create a SNP-gene pair, merely for easier description, as proximity-based assignments may overlook distal regulatory effects. In detail, notable SNPs along with its corresponding nearest gene in the 1^st^ SNP category, “Severe COVID-19-Specific SNPs,” include rs10774679 (*OAS3*), rs17169628 (*SLC22A31*), rs12329760 (*TMPRSS2*), rs12660421 (*FOXP4*), rs17078348 (*SLC6A20*), rs190509934 (*ACE2*), rs2834164 (*IFNAR2*), rs3785884 (*MAPT*), rs7251000 (*DPP9*), and rs76608815 (*SLC5A3*), among others (Table 1), and this SNP category contains a total of 27 SNPs showing stronger associations with severe COVID-19 than with mild cases (Fig. 2 and Fig. S2). Meanwhile, for the 2^nd^ SNP category, “SNPs Associated with Both Severe and Mild COVID-19”, five out of 15 SNP-gene pairs, including rs2290859 (*NXPE3*), rs505922 (*ABO*), rs149533170 (*IFNA21*), rs34536443 (*TYK2*), and rs9916158 (*GSDMA/B*), demonstrate consistent associations across GWAS datasets without differential effects between hospitalized and non-hospitalized groups (Fig. 2 and Fig. S3). Notably, *ABO* (rs505922) has been reported as a genetic locus associated with long COVID[5]. Several other immune-related genes were found close to other candidate SNPs, including *IFNA21* (rs149533170), *TYK2* (rs34536443), *GSDMA/B* (rs9916158), *BCL11A* (rs1123573), *ELF5* (rs2924480), and *JAK1* (rs11208552), in accordance with the role of immune dysregulation in both hospitalized and non-hospitalized COVID-19[19–24]. Additionally, *FUT2* is found closely with rs492602 and encodes fucosyltransferase 2, a key enzyme in the ABO antigen synthesis pathway, further underscoring the involvement of *ABO* in COVID-19 outcomes[25]. Additionally, four genes—*TAC4*[26], *NXPE3[27]*, *NSF*[28], and *FOXG1*[29]—linked to neuronal functions were close to rs75432325, rs2290859, rs1378358, and rs150497803, respectively, while three genes—*ZKSCAN1*[30], *BCL11A*, and *FOXG1*—identified as transcription factors were linked to rs2897075, rs1123573, and rs150497803, respectively.

We also evaluated sex-biased associations for these 42 candidate SNPs in the first 2 SNP categories (Fig. S4). Due to the smaller sample sizes in the sex-stratified COVID-19 hospitalization GWASs from UK Biobank, only 15 SNPs reached nominal significance (*P <* 0.05) in at least one of these three GWASs, including COVID-19 hospitalization for males and females, and differential COVID-19 hospitalization between males and females. Of these, rs17078348 (*SLC6A20*), a well-established severe COVID-19 risk SNP[31], showed nominal significance in both male and female cohorts of COVID-19 hospitalization. Five SNPs displayed nominal associations exclusively in females, including rs75432325 (*TAC4*), rs505922 (*ABO*), rs12660421 (*FOXP4*), rs7251000 (*DPP9*), and rs11766643 (*TFR2*). Eight SNPs were nominally associated only in males, including rs897075 (*ZKSCAN1*), rs117169628 (*SLC22A31*), rs7515509 (*AK5*), rs76608815 (*SLC5A3*), rs12585036 (*ATP11A*), rs5023077 (*FBRSL1*), rs10774679 (*IFNAR21*), and rs12329760 (*HLA-C*). None of these SNPs passed the stringent multiple testing correction threshold (*P <* 6×10^-4^), and local Manhattan plots for 12 SNPs revealed relatively weaker association signals in the sex-stratified GWAS in contrast with the stronger signals observed in HGI-B2 (Fig. S5). In summary, sex-biased GWAS analysis did not identify SNPs with statistically significant sex-biased associations with COVID-19 hospitalization, with only 15 SNPs with nominal significance, such as rs505922 (*ABO*) and rs12660421 (*FOXP4*).

### Candidate SNPs Specifically Associated with Non-Hospitalized COVID-19

To identify potential candidate SNPs specific to non-hospitalized COVID-19 cases, we screened for SNPs that showed suggestive significant association signals in HGI-B1 (hospitalized vs. non-hospitalized, *P <* 1×10^-5^) and HGI-B1-vs-B2 (differential GWAS between HGI-B1 and HGI-B2, *P <* 0.05) but were non-significant in HGI-B2 (hospitalized vs. general population, *P* > 0.05) and were not in proximity to any genome-wide significant SNPs in HGI-B2. We also only focused on top hits with MAF > 0.01 in the HGI-C2 with the largest sample size compared to other COVID-19 GWASs from HGI. This selection prioritized 20 independent SNPs potentially specifically associated with mild (non-hospitalized) COVID-19 cases (Table 2; Fig. S6), and only 2 out of them have nominal association of P < 0.05 in HGI-G2, suggesting most of them are not COVID-19 susceptibility related variants. All 20 SNPs showed suggestive differential effect sizes between the two GWASs of HGI-B1 and HGI-B2 (*P <* 1×10^-5^). Interesting findings were observed by linking these candidate SNPs with their adjacent genes. For example, rs62401842 (*KCTD16*) exhibited the strongest association in HGI-B1 (*P =* 8.53×10^-7^), while showing no association in HGI-B2 (*P =* 0.38), resulting in a significant difference between HGI-B1 and HGI-B2 (*P =* 3.18×10^-7^). *KCTD16*, encoding potassium channel tetramerization domain-containing 16[32], is highly expressed in brain tissues according to the GTEx database[17]. In addition, eight SNPs illustrated in Fig. S6 were mapped to genes with brain-related functions, including rs9799354 (*NLGN1*[4]), rs112842080 (*CPLX2*[33]), rs61858037 (*NRG3*[34]), rs61939166 (*KIF21A*[35]), rs367777 (*NAV3*[36]), rs56143829 (*WASF3*[37]), rs11454577 (*AKAP6*[38]), and rs6049828 (*SYNDIG1*[39]). Among these, *WASF3* has been implicated in mitochondrial dysfunction and may mediate exercise intolerance in myalgic encephalomyelitis/chronic fatigue syndrome[37], a phenotype highly similar to long COVID. Additionally, among these 20 candidates SNPs identified, only three SNPs (rs62401842 [*KCTD16*], rs9799354 [*NLGN1*], and rs112842080 [*CPLX2*]) exhibit male-biased associations with COVID-19 hospitalization at nominal significance levels (Fig. S7). Furthermore, six candidate SNPs, including rs62401842 (*KCTD16*), rs9799354 (*NLGN1*), rs112842080 (*CPLX2*), rs56143829 (*WASF3*), rs4737438 (*PENK*), and rs6049828 (*SYNDIG1*), demonstrate sex-biased association patterns within a genomic window ranging from 500kb to 1000kb harboring other SNPs displaying nominal association signals to COVID-19 hospitalization (Fig. S7). However, the limited sample sizes in sex-stratified COVID-19 GWAS preclude any candidate SNPs from achieving statistical significance after correction for multiple testing in analyses of sex-biased associations with COVID-19 hospitalization. In summary, while no genome-wide significant SNPs specific to non-hospitalized COVID-19 cases were identified, these 20 SNPs along with their adjacent genes provide suggestive evidence of association with mild COVID-19, of which 18 are not associated with COVID-19 susceptibility.

**Table 2.** Candidate SNPs specifically associated with non-hospitalized COVID-19 cases.

| Variant information |  |  |  |  |  | HGI-B1 |  |  | HGI-B2 |  |  | HGI-C2 |  |  | HGI-B1-vs-B2 |
| --- | --- | --- | --- | --- | --- | --- | --- | --- | --- | --- | --- | --- | --- | --- | --- |
| chr | pos | rsid | allele | AF | gene | beta | se | p | beta | se | p | beta | se | p | DiffPval |
| 3 | 174147743 | rs9799354 | C | 0.556 | *NLGN1* | 0.08 | 0.02 | 5.38E-06 | 0.01 | 0.01 | 2.71E-01 | -0.01 | 0.00 | 1.79E-02 | 6.21E-06 |
| 5 | 128241667 | rs201484359 | G | 0.304 | *SLC27A6* | 0.13 | 0.03 | 8.55E-06 | 0.02 | 0.01 | 1.25E-01 | -0.01 | 0.01 | 8.91E-02 | 2.58E-05 |
| 5 | 143904835 | rs62401842 | A | 0.042 | *KCTD16* | 0.27 | 0.06 | 8.53E-07 | 0.02 | 0.03 | 3.82E-01 | 0.00 | 0.01 | 8.72E-01 | 3.18E-07 |
| 5 | 175248251 | rs112842080 | T | 0.021 | *CPLX2* | -0.36 | 0.07 | 1.58E-06 | -0.04 | 0.03 | 2.53E-01 | 0.03 | 0.02 | 6.69E-02 | 1.25E-06 |
| 6 | 37086436 | rs147171940 | A | 0.038 | *PIM1* | -0.19 | 0.04 | 5.96E-06 | -0.04 | 0.02 | 1.33E-01 | 0.01 | 0.01 | 5.74E-01 | 5.92E-05 |
| 6 | 151444801 | rs76392050 | T | 0.010 | *MTHFD1L* | 0.70 | 0.16 | 6.10E-06 | 0.10 | 0.06 | 6.16E-02 | 0.03 | 0.03 | 3.84E-01 | 5.88E-06 |
| 8 | 57438048 | rs4737438 | A | 0.378 | *PENK* | 0.08 | 0.02 | 2.97E-06 | 0.01 | 0.01 | 2.79E-01 | 0.00 | 0.00 | 5.30E-01 | 2.98E-06 |
| 8 | 68035003 | rs148340257 | T | 0.065 | *CSPP1* | -0.22 | 0.05 | 8.05E-06 | -0.03 | 0.02 | 1.77E-01 | 0.00 | 0.01 | 8.82E-01 | 7.21E-06 |
| 9 | 123344199 | rs10760104 | T | 0.819 | *CDK5RAP2* | -0.13 | 0.03 | 2.37E-06 | -0.01 | 0.01 | 4.57E-01 | 0.00 | 0.01 | 8.77E-01 | 5.36E-07 |
| 10 | 54976003 | rs55654117 | C | 0.272 | *MBL2* | 0.10 | 0.02 | 1.85E-06 | 0.01 | 0.01 | 2.61E-01 | 0.00 | 0.00 | 4.50E-01 | 1.13E-06 |
| 10 | 84981211 | rs61858037 | T | 0.091 | *NRG3* | 0.15 | 0.03 | 9.21E-06 | 0.02 | 0.02 | 1.62E-01 | 0.00 | 0.01 | 9.58E-01 | 2.31E-05 |
| 12 | 39502966 | rs61939166 | G | 0.011 | *KIF21A* | -0.40 | 0.09 | 3.60E-06 | -0.03 | 0.04 | 5.36E-01 | -0.02 | 0.02 | 3.63E-01 | 1.10E-06 |
| 12 | 78552292 | rs367777 | T | 0.358 | *NAV3* | -0.10 | 0.02 | 3.13E-06 | -0.01 | 0.01 | 2.77E-01 | 0.01 | 0.00 | 2.39E-01 | 1.89E-06 |
| 13 | 27062460 | rs56143829 | T | 0.025 | *WASF3* | 0.24 | 0.05 | 1.26E-06 | 0.01 | 0.03 | 6.03E-01 | 0.00 | 0.01 | 7.56E-01 | 3.23E-07 |
| 13 | 63988259 | rs9570861 | A | 0.552 | *PCDH20* | 0.08 | 0.02 | 5.51E-06 | 0.02 | 0.01 | 1.07E-01 | -0.01 | 0.00 | 4.06E-02 | 4.83E-05 |
| 14 | 32937383 | rs11454577 | G | 0.589 | *AKAP6* | -0.09 | 0.02 | 5.54E-06 | -0.01 | 0.01 | 3.22E-01 | 0.00 | 0.01 | 6.24E-01 | 7.71E-06 |
| 16 | 84887921 | rs113067468 | G | 0.030 | *CRISPLD2* | -0.30 | 0.07 | 4.48E-06 | -0.05 | 0.03 | 1.20E-01 | 0.00 | 0.02 | 8.21E-01 | 2.18E-05 |
| 17 | 3454928 | rs115744301 | G | 0.035 | *TRPV3* | 0.66 | 0.15 | 5.21E-06 | 0.02 | 0.05 | 6.89E-01 | -0.03 | 0.03 | 2.52E-01 | 2.53E-07 |
| 17 | 13775336 | rs11869231 | T | 0.310 | *COX10* | -0.11 | 0.02 | 2.38E-06 | 0.00 | 0.01 | 6.52E-01 | 0.00 | 0.00 | 9.85E-01 | 2.26E-07 |
| 20 | 24627964 | rs6049828 | C | 0.567 | *SYNDIG1* | 0.09 | 0.02 | 7.13E-06 | 0.01 | 0.01 | 2.65E-01 | 0.00 | 0.00 | 3.51E-01 | 5.43E-06 |

Note: abbreviations in the table headers are similar to that of Table 1.

### Evaluating Associations of 62 Prioritized Candidates SNPs with Long COVID

To investigate the associations of these aforementioned 62 candidates SNPs for future investigation of long COVID, the four published long COVID GWASs from HGI were used to examine association signals for them. Notably, only one candidate SNP, rs12660421 of *FOXP4*, passed the traditional genome-wide significance threshold and significantly associated with long COVID (*P =* 1×10^-9^) in one of four long COVID GWASs, i.e. LongCOVID-N2 GWAS (strict cases of long COVID after test-verified SARS-CoV-2 infection [*n* = 3,018] vs. general population controls [*n* = 994,582]) (Table S1, Fig. S8-S10). Among other 3 alternative long COVID GWASs with slightly different case-control designs for long COVID, including LongCOVID-W2, LongCOVID-N1, and LongCOVID-W1 (see method for detailed case-control designs for them), rs12660421 (*FOXP4*) is also nominally significantly associated with long COVID, with *P*-values equal to 9.7×10^-7^, 4.3×10^-3^, and 4.95×10^-2^, respectively. However, rs12660421 (*FOXP4*) tends to be more strongly associated with COVID susceptibility (*P =* 7.4×10^-15^) and hospitalization (*P =* 2.9×10^-10^), supporting the SNP’s association with multiple COVID-19 and long COVID phenotypes. For another 61 remaining candidate SNPs, there are 8 SNPs, comprising rs505922 (*ABO*), rs7660881 (*SLC5A3*), rs6186040 (*SFTPD*), rs2834164 (*IFNAR2*), rs1077467 (*OAS3*), rs2290859 (*NXPE3*), rs1707834 (*SLC6A20*) and rs1128420 (*CPLX2*), showing nominal significance in at least one long COVID GWAS, all of which displaying much weaker association signals compared to that from acute-phase COVID-19 GWASs (Table 1 and 2, Table S1; Fig. S8-S10). Additionally, 43 out of 62 candidate SNPs, including *ELF5* (rs141160516), *TAC4* (rs28704677), *FUT2* (rs557806), *RAB2A* (rs113059174), *ABO* (rs544873), *GSDMA*/*B* (rs72832927), *HLA*-C (rs3218825), *TYK2* (rs8112028), *OAS3* (rs72487538), *KCTD16* (rs114210747), *WASF3* (rs75947067), *ATP11A* (rs3832907), *PCDH20* (rs34409482), *SLC27A6* (rs704234), and others (see Table S2), have at least one adjacent SNP displaying long COVID association signals with *P <* 0.001, indicating that these loci are warranted for further investigation in independent long COVID samples (Table S2 and Fig. S10). Taken together, we observed much weaker associations of these prioritized SNPs as well as its adjacent SNPs with long COVID.

### Evaluating Other Published Long COVID SNPs in Current COVID-19 GWASs

Previously published long COVID risk SNPs reported by Taylor et al.[4] and Chaudhary et al.[5] were investigated among three acute-phase COVID-19 GWASs of HGI (Table S3). The rationale is based on the hypothesis that larger sample sizes and higher statistical power of the HGI datasets, such as severe COVID-19 and susceptibility to SARS-CoV-2 infection, have enough power to prioritize candidate loci related to long COVID. Moreover, overlap between our prioritized long COVID SNPs and those reported in previous studies would aid for future in-depth investigation on long COVID.

Among the three SNPs (rs643434, rs2080090, and rs9273363) reported by Chaudhary et al.[5] (Fig. S11 and Table S3), rs643434 (*ABO*) and rs2080090 (*BPTF*) demonstrated significant associations with both COVID-19 hospitalization (HGI-B2) and susceptibility to COVID-19 (HGI-C2). In detail, rs643434 (*ABO*) surpassed genome-wide significance thresholds in both HGI-B2 (hospitalized vs. general population; *P =* 6.7×10^-24^) and HGI-C2 (COVID19 infected vs. general population; *P =* 8.7×10^-85^) and showed non-significant association in HGI-B1 (hospitalized vs. non-hospitalized COVID-19; *P =* 0.6) but demonstrated significantly differential effect sizes (*P =* 6×10^-5^) between HGI-B1 and HGI-B2. This supported its classification as a “category 2” COVID-19 SNP (associated with both severe and mild COVID-19 phenotypes). Similarly, rs2080090 (*BPTF*) showed stronger associations in HGI-B2 (*P =* 1.9×10^-4^) than in HGI-C2 (*P =* 2.5×10^-2^), with no significant association in HGI-B1. This SNP was also categorized as a suggestive “category 2” COVID-19 SNP. In contrast, rs9273363 (*HLA-DQB1*) did not exhibit significant associations across any of the HGI GWAS datasets analyzed (all *P*s > 0.2). Nevertheless, among these 62 candidate SNPs, rs17219281 (*HLA-DQA1*), a “category 1” COVID-19 SNP that tends to be more likely associated with hospitalized rather than mild COVID-19 (see Table 1), is ∼50kb downstream of rs9273363 (*HLA-DQB1*), strongly implicating that rs17219281 may be a promising candidate SNP related to both COVID-19 hospitalization and long COVID. Taken together, the three recently published long COVID loci, including *ABO*, *BPTF*, and *HLA-DQB1*, were completely covered by these 62 prioritized SNPs and shown associations with either COVID-19 hospitalization or both COVID-19 hospitalization and non-hospitalization.

Meanwhile, of the 80 SNPs reported by Taylor et al.[4], 12 displayed nominally significant associations in at least one HGI GWAS dataset (Table S3 and Fig. S11), including four SNPs associated with HGI-B2, five with HGI-B1, and seven with HGI-C2. Notably, rs4303401 (*FRMD6*) surpassed the multiple testing correction threshold (*P <* 1.5×10^-4^), representing a strong candidate SNP. Further explorative analysis by scanning a ±500kb genomic window around the reported SNPs identified additional suggestive genome-wide significant associations in the HGI datasets. For example, in Fig. S11, rs62401842 (proximal to *KCTD16*, a brain-biased gene) and rs7671107 and rs1017716 (a GTEx eQTL of *ANAPC4*, a gene implicated in aging) emerged as promising candidates. Other noteworthy SNPs in Fig. S11 include rs12602210 (*BPTF*), rs17412601 and rs2290859 (both mapped to *NXPE3*), demonstrating suggestive or genome-wide significance in at least one HGI GWAS dataset. In summary, four loci, including *KCTD16*, *ANAPC4*, *BPTF*, and *NXPE3*, revealed by Taylor et al[4]. derived from SNP network analysis are independently confirmed to be nominally associated with COVID-19 related phenotypes in the current study; *BPTF* is a locus confirmed to be associated with long COVID by Chaudhary et al., Taylor et al., and also associated with acute-phase COVID-19 in current study.

#### Transcriptome-Wide Association Study Identifies Candidate Genes Adjacent to Prioritized SNPs

To investigate COVID-19 candidate SNPs and its adjacent genes, we further performed a transcriptome-wide association study (TWAS) using S-MetaXcan across 48 GTEx tissues by correlating genotype-imputed gene expression with acute-phase COVID-19 and long COVID. Although this simple analysis is not comprehensive as that of colocalization or fine-mapping, it would be a first step to explore potential candidate genes that would be investigated deeply with colocalization or fine-mapping in the future. We first utilized COVID-19 GWAS summary statistics from 4 HGI GWASs: HGI-B1, HGI-B2, HGI-C2, and HGI-B1-vs-B2. The analysis targeted 62 candidate SNPs previously prioritized and aimed to identify their adjacent genes whose expression levels may be associated with COVID-19 hospitalization or susceptibility phenotypes, and then 4 long COVID GWASs with slightly different definitions in terms of cases and controls were performed for TWAS using the same method.

We evaluated whether these candidate SNPs and its adjacent genes were covered by TWASs of acute-phase COVID-19. Of the 62 candidate SNPs, two SNPs (rs190509934 near *ACE2* and rs9570861 near *OR7E156P*) were not included in the TWAS analysis. The former resides on chromosome X and was excluded by S-MetaXcan because the software only focuses on autosomal genes, while the latter SNP has no protein-coding genes within a 500-kb window where the SNPs is located at the center. Although there are 879 adjacent genes to these 62 candidate SNPs, only 776 out of them are corresponding to the remaining 60 SNPs that were testable by these COVID TWASs (Fig. S12) with detectable expression across 48 GTEx tissues.

Next, to simplify the interpretation for the TWAS results, we retained only the strongest association (smallest p-value) for each gene across 48 GTEx tissues. This approach was justified by the hypothesis that specific tissues, rather than all tissues, are likely to drive the pathogenicity of different COVID phenotypes. Applying varying significance thresholds (*P <* 0.05, 0.01, 0.001, 0.0001, 0.00001), the analysis revealed 422, 152, 43, 19, and 11 adjacent genes meeting these criteria in at least one of the four HGI datasets (Fig. S12, Table S4). Using a genome-wide significance threshold (*P <* 3.5×10^-7^), we found that top hits are originated exclusively from COVID-19 hospitalization and susceptibility GWAS, with no genes passed this threshold in long COVID TWASs (Fig. 3). The top three loci displayed significant associations are: (1) *DPP9*, showing the strongest association signal (*P =* 4.6×10^-18^) in adipose visceral omentum tissue; (2) *CXCR6* gene cluster, which included multiple genes (*CXCR6*, *CCR3*, *CCR1*, and *LZTFL1*), displaying the most significant association (*P =* 1.3×10^-10^) in nerve tibial tissue; (3) *MUC1* gene cluster, comprising *MUC1*, *THBS3* and *FAM189B*, highlighted in multiple GTEx tissues, including lung and colon. Additional loci with suggestive TWAS significance (*P <* 3.5×10^-6^) included *FYCO1*, *SLC6A20*, and *XCR1*, specifically showing associations in HGI GWASs B1, B2, and C2 among distinct GTEx tissues.

Further determination revealed that these candidate genes proximal to 60 prioritized SNPs accounted for all top TWAS signals across the eight HGI TWASs, with the exception of *CCR5*, *EXOSC7*, *FLT1P1*, *RLN1*, *GBAP1*, *ARHGEF2*, *ZSCAN31*, *ZKSCAN3*, *PGMSP2*, *PAQR5*, and *TMIE* (Fig. S13A). Further in-depth evaluation of these outlier genes in long COVID TWASs revealed most of them were insignificant (Fig. S13B and S14), supporting the feasibility of only focusing on these top candidate SNPs and their adjacent genes for future investigation on long COVID.

For explorative analysis, we then further focused on the associations of these adjacent genes in long COVID TWASs across all 48 GTEx tissues using a relatively weaker association threshold, i.e., the nominal significance threshold of *P <* 0.05, among these top COVID-19 TWAS genes (Table 3). In details, *THBS3* is the gene showing ubiquitous nominal association with long COVID among multiple GTEx tissues, such as brain cerebellum, nerve tibial, spleen, adrenal gland, colon sigmoid, testis and esophagus mucosa; *CCR3* is observed to be nominally correlated with long COVID in the GTEx tissue heart atrial appendage; *MUC1* is predicted to be nominally associated with long COVID in the GTEx tissue colon transverse, and *DPP9* expression associates with long COVID in both GTEx tissues colon transverse and testis. Furthermore, using a relaxed threshold (*P <* 0.05), we prioritized additional adjacent genes potentially related to long COVID (Table 3 and Fig. S15-16). These included *OAS3* (rs10774679), *TMPRSS2* (rs12329760), *HLA-A* (rs4s9260038), *HLA-C* (rs1634761), *HLA-DQA1* (rs17219281), *WASF3* (rs65143829), *IFNAR2* (rs2834164), *SFTPD* (rs61860402), and *GSDMA/B* (rs9916158). Notably, the well-established *ABO* locus, a COVID-19 susceptibility locus, did not show significant TWAS signals.

**Table 3.**
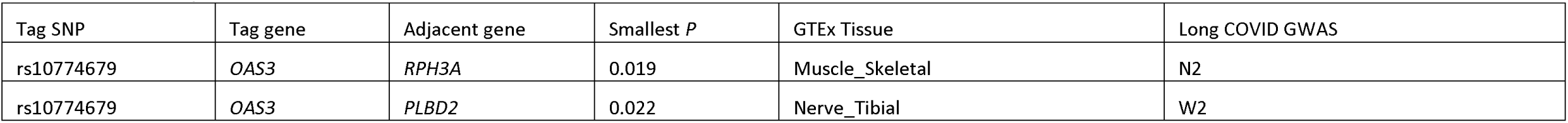
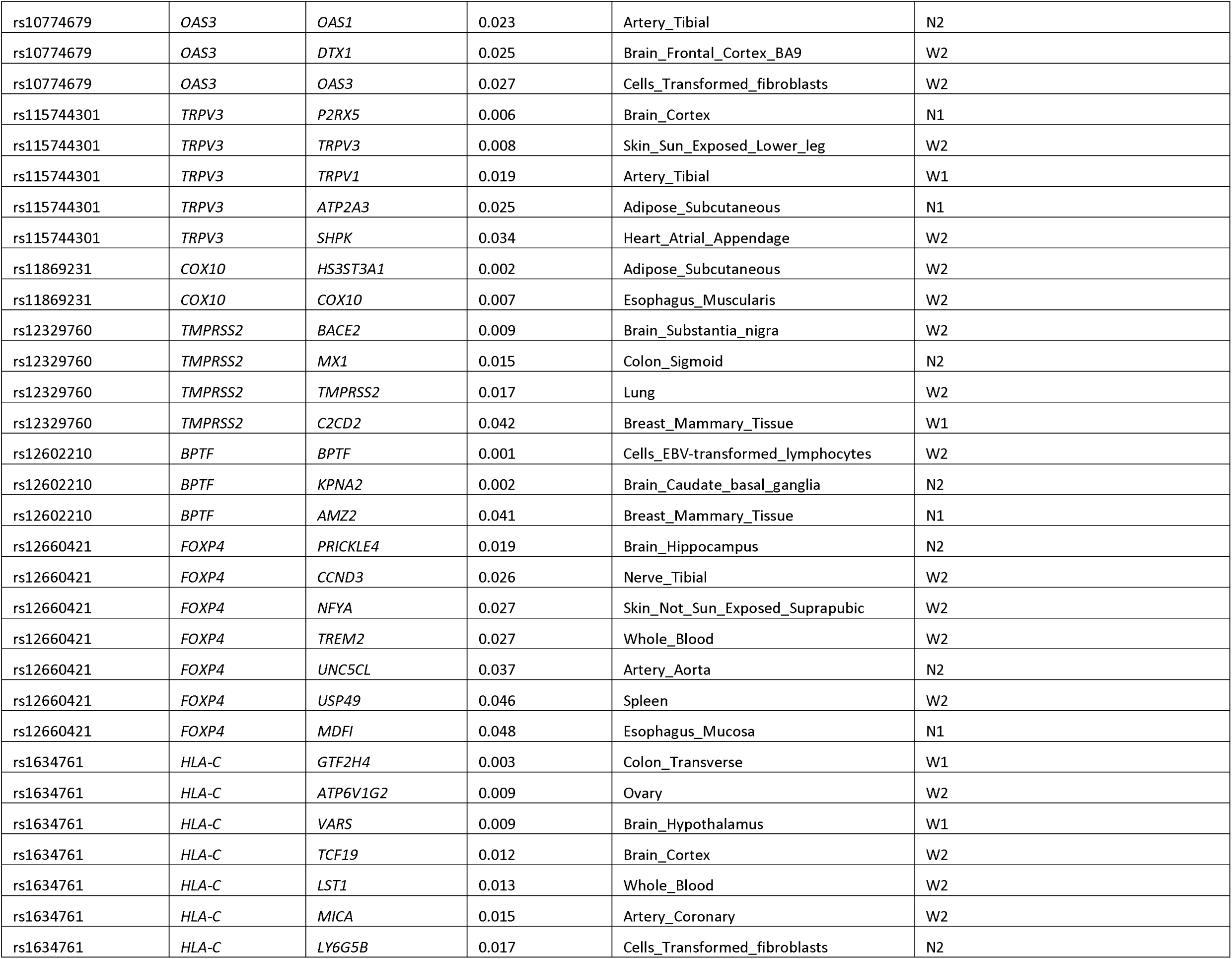
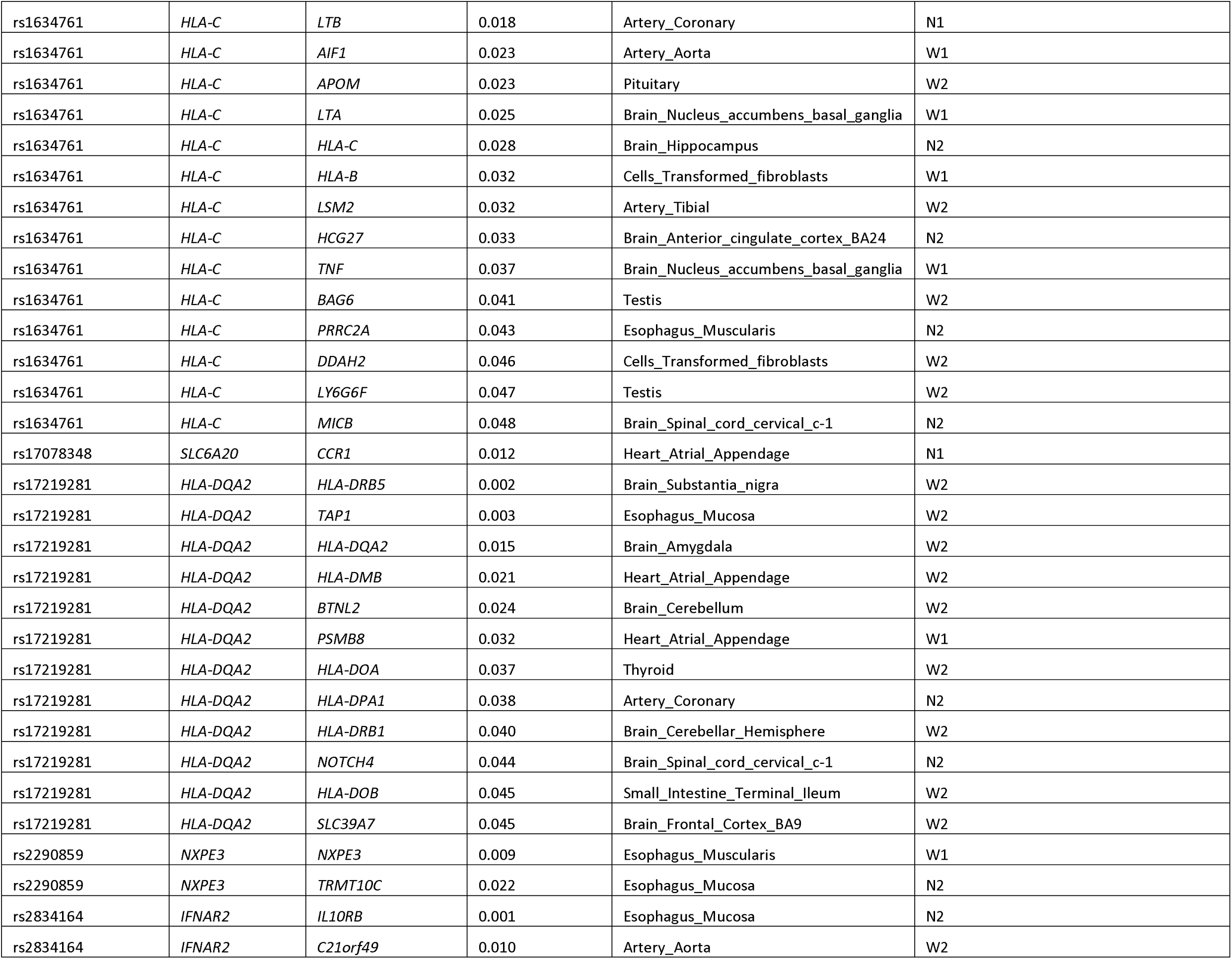
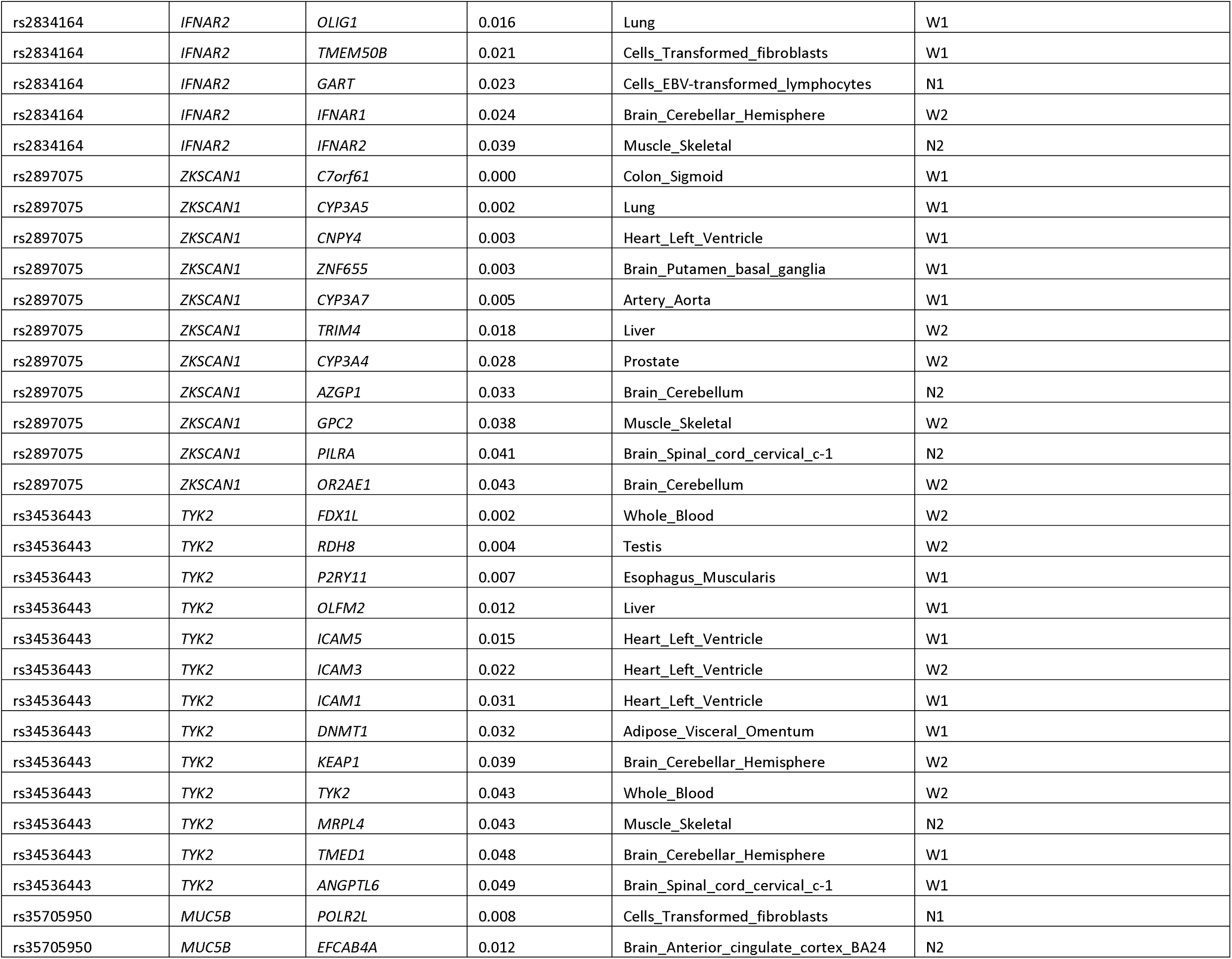
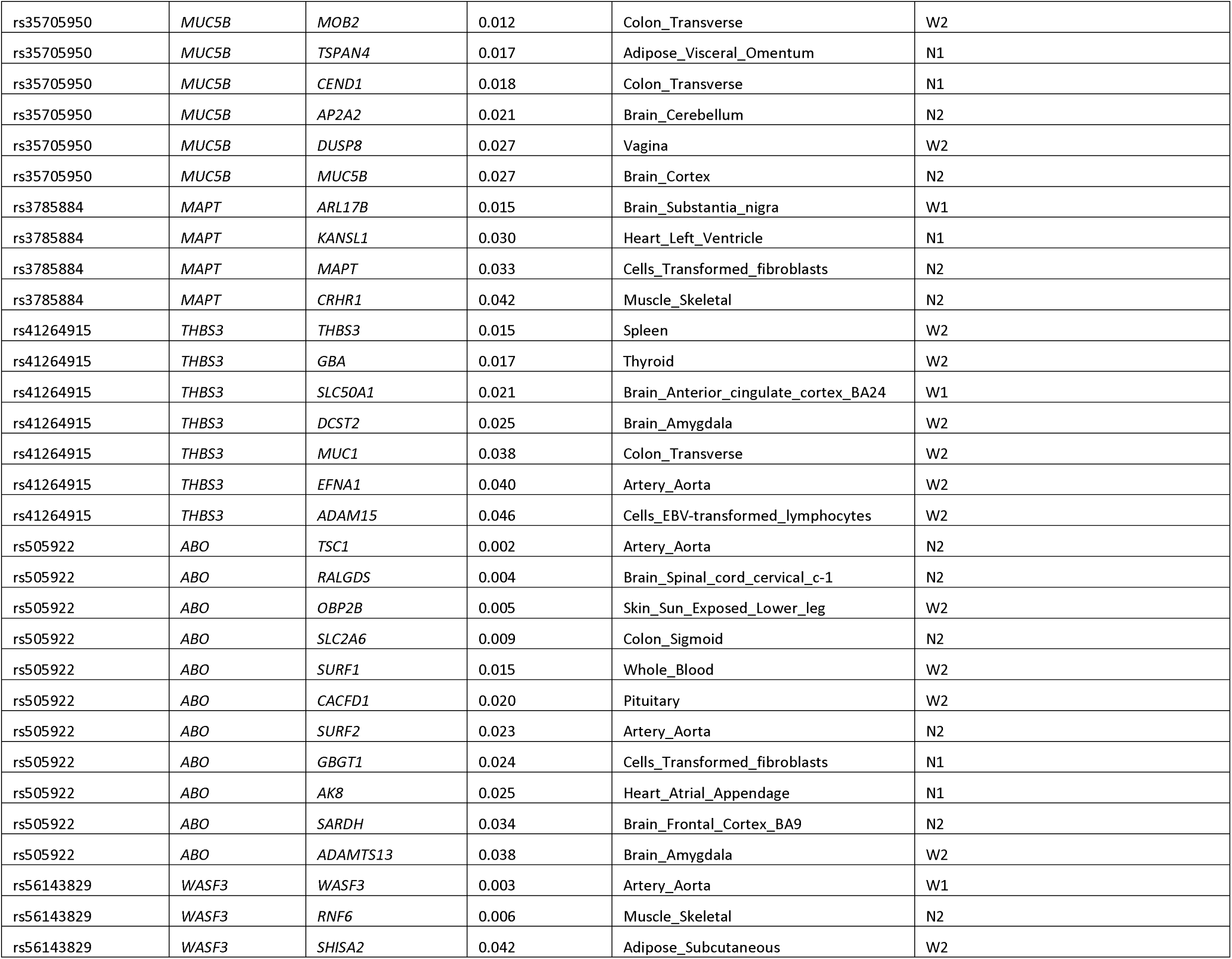
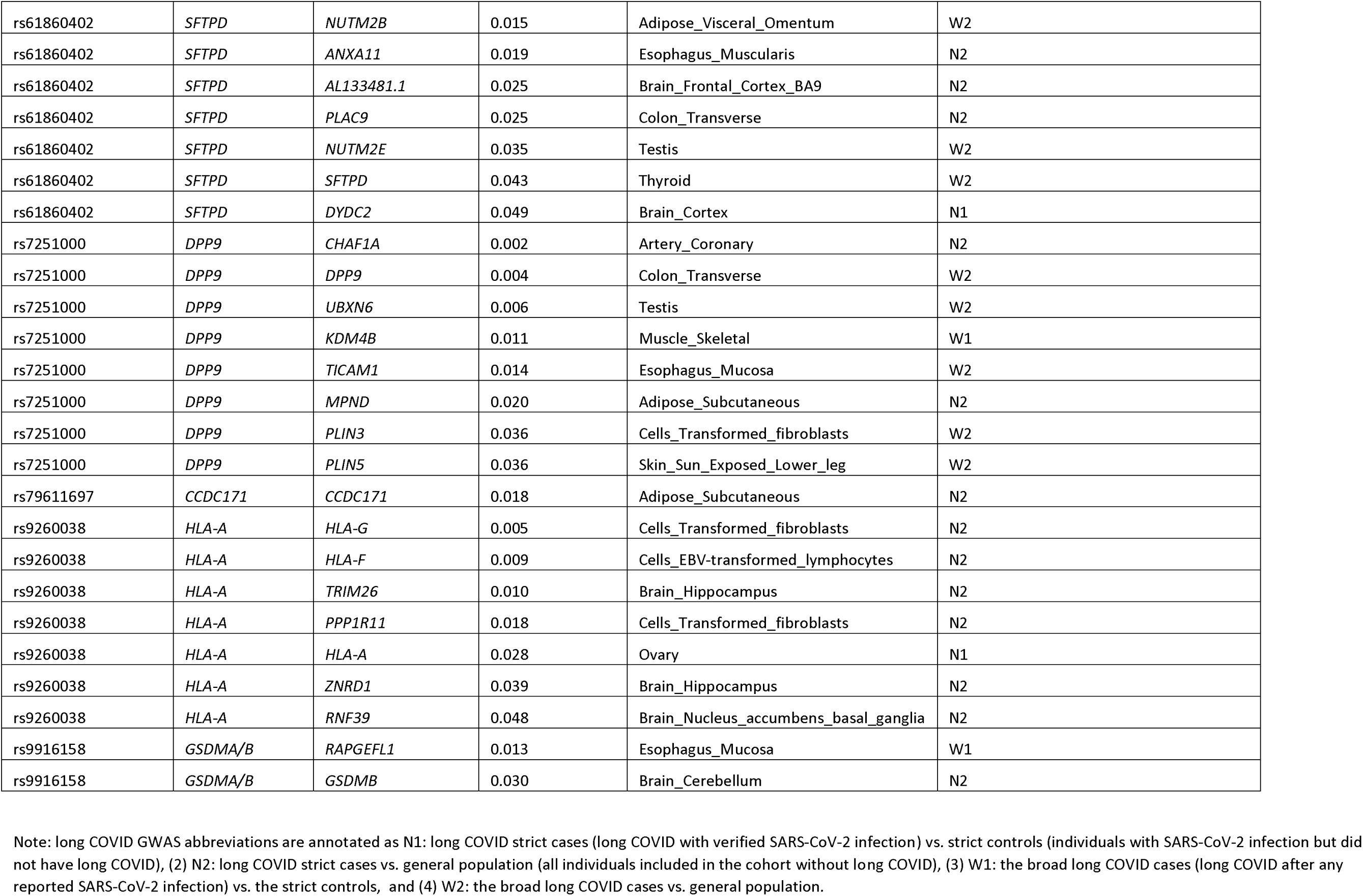
Top genes adjacent to prioritized candidate SNPs (n=23) in long COVID related TWASs, with the smallest *P across 48 GTEx tissues displayed*.

In summary, TWAS analyses revealed much weaker associations (only some of them passed the nominal significance *P* < 0.05) for 52 SNP-gene pairs with long COVID, with genes such as *DPP9, CCR1*, *THBS3*, and *MUC1* prioritized as potential targets for both acute and long COVID phenotypes.

### Phenome-Wide Analysis of Genes Expression Association

To investigate gene expression associations with acute-phase COVID-19 and long COVID-related phenotypes, we also performed a phenome-wide analysis of gene expression associations (Pheno-TWAS) analysis targeting adjacent genes located within a 1Mb-window of 62 candidate SNPs. Of the 918 tested TWAS genes, 400 genes (43.6%), including 87 *HLA* loci (e.g., *HLA-DQA2*, *HLA-A*, and *HLA-C*; Fig. S17-19), passed a stringent bonferroni-adjusted significance threshold (*P <* 2.7×10^-9^). Genes located near *HLA* loci emerged as top hits, demonstrating significant associations with immune-related disorders such as asthma, rheumatoid arthritis, psoriasis, coeliac disease, and systemic lupus erythematosus—conditions that may be risk factors for acute COVID-19 or long COVID. In addition to *HLA*-associated loci, other 313 genes across 44 non-*HLA* loci surpassed the genome-wide significance threshold in TWASs. Key findings include: (1) *IFNAR2*: strongly associated with COVID-19 hospitalization (*P* = 2.6×10^-10^; Fig. S20); (2) *GSDMB*: linked to asthma (*P* = 2.7×10^-57^; Fig. S21); (3) *BPTF*: associated with trunk fat percentage, a key marker for metabolic health (*P* = 1×10^-18^; Fig. S22); (4) *CCDC171*: associated with body fat percentage, a key indicator of fitness and health (*P* = 2×10^-35^; Fig. S23). Interestingly, *FOXP4*, which did not exhibit significant associations with long COVID phenotypes in prior TWASs, demonstrated a suggestive association with sleep duration (*P* = 1.1×10^-5^; Fig. S24), and sleeping less than 7 hours is suggested to be linked with obesity, diabetes, and heart disease. In summary, Pheno-TWAS identified 45 candidate SNPs associated with genome-wide significant expression-linked phenotypes. These include three major *HLA* loci (*HLA-DQA2*, *HLA-A*, and *HLA-C*) as well as non-*HLA* loci such as *IFNAR2*, *BPTF*, *CCDC171*, and *GSDMB*. The implicated phenotypes, ranging from COVID-19 hospitalization and immune disorders to body composition traits and sleep duration, provide a broad spectrum of potential biological pathways linking to acute COVID-19 or long COVID pathophysiology. Top results, including nominally significant associations, are provided in Table S6.

## DISCUSSION

Long COVID, characterized by over 200 symptoms spanning multiple organ systems, presents significant challenges for GWAS due to difficulties in assembling large, representative long COVID samples. To date, only four genome-wide significant loci, including *FOXP4*, *ABO*, *HLA-DQA1*, and *BPTF*, have been identified to be genome-wide significantly associated with long COVID by different studies[5, 14]. Our integrative analysis prioritized 62 candidate loci for future investigation on long COVID based on their associations with acute-phase COVID-19 phenotypes, such as COVID-19 susceptibility, hospitalization, and non-hospitalization. These candidate SNPs were obtained by leveraging large-scale COVID-19 GWAS datasets from the HGI. Our prioritization would facilitate future investigation on long COVID as currently published long COVID GWASs have the inherent limitations of limited sample sizes and phenotype heterogeneity, and SARS-CoV-2 susceptibility and COVID-19 hospitalization can be used as proxy phenotypes to study long COVID in the future. By categorizing genetic variants into three groups, covering those associated with hospitalized COVID-19, those linked to both hospitalized and non-hospitalized cases, and those specific to non-hospitalized cases, we prioritized potential SNPs that might be associated with long COVID.

There are 42 SNPs prioritized to be associated with either COVID-19 hospitalization or both COVID-19 hospitalization and non-hospitalization, including well-documented variants such as rs12660421 (*FOXP4*) and rs505922 (*ABO*), which have been implicated in severe COVID-19 outcomes and long COVID susceptibility. Notably, additional loci like rs17219281 (*HLA-DQA1*), rs9260038 (*HLA-A*), rs1634761 (*HLA-C*), rs7251000 (*DPP9*), rs9916158 (*GSDMB*), rs12602210 (*BPTF*), rs17078348 (*CCR1*), and rs41264915 (*THBS3*), rs190509934 (*ACE2*) and rs12329760 (*TMPRSS2*) were selected as candidate loci. Furthermore, a unique category comprising 20 SNPs emerged specifically in non-hospitalized COVID-19 cases. These include rs62401842 (*KCTD16*) and rs56143829 (*WASF3*), mapped to genes involved in brain-related functions. However, future investigations are warranted for these candidate SNP-gene pairs in long COVID.

One important consideration in interpreting our results is the conceptual distinction among the HGI-B1, HGI-B2, and HGI-C2 phenotypes. While HGI-C2 predominantly reflects susceptibility to SARS-CoV-2 infection and HGI-B1 isolates disease severity among infected individuals, HGI-B2 represents a composite phenotype that incorporates both infection susceptibility and severe disease progression because hospitalized COVID-19 cases are compared against the general population. Consequently, loci identified in HGI-B2 cannot be interpreted solely as severity-associated signals, as these associations may arise from multiple biological mechanisms, including increased susceptibility to infection, increased likelihood of hospitalization following infection, or both. This mixed phenotype structure motivated our integrative comparison framework across the three GWASs. By leveraging the different control definitions between HGI-B1 and HGI-B2, we aimed to partially disentangle genetic contributions associated with severe versus mild COVID-19 phenotypes through the HGI-B1-vs-B2 differential GWAS. However, because HGI-B2 uses the general population as controls, differential association signals between HGI-B1 and HGI-B2 could theoretically also reflect susceptibility-related effects. Nevertheless, the empirical association patterns observed in the prioritized loci (Table 2) argue against susceptibility being the dominant explanation for most candidate SNPs. Specifically, the majority of prioritized mild-COVID-19 loci demonstrated significant association signals in HGI-B1 but weak or non-significant associations HGI-C2. Because HGI-C2 is primarily designed to capture infection susceptibility, the absence of strong HGI-C2 signals suggests that these loci are less likely to represent generalized susceptibility variants and instead supports the interpretation that the differential GWAS framework enriches for loci associated with non-hospitalized or milder COVID-19 phenotypes. Nonetheless, complete separation of susceptibility, severity, and mild-disease effects is not possible using current HGI datasets alone. Therefore, the SNP categories identified in this study should be interpreted as operational classifications derived from comparative GWAS architectures rather than strictly isolated biological pathways, and future GWASs with more refined clinical phenotyping, particularly for non-hospitalized and longitudinal COVID-19 populations, will be important for validating these distinctions.

In conclusion, our findings underscore the utility of integrative approaches to prioritize genetic risk loci for future investigation of long COVID. The prioritization of loci such as *FOXP*4, *ABO*, and *HLA-DQA1*, as well as *BPTF*, highlights potentially shared biological pathways between acute COVID-19 severity and its chronic manifestations, such as long COVID. Additionally, the identification of brain-enriched loci in non-hospitalized cases aligns with emerging evidence linking long COVID to neuroinflammatory and neurodegenerative processes. Future studies, particularly those incorporating diverse populations and long-term follow-up data, are essential to validate these associations and unravel the intricate mechanisms underlying long COVID.

## LIMITATIONS

Certain limitations warrant consideration in our study. First, the reliance on proxy phenotypes may obscure SNPs specific to mild COVID-19 cases, due to the skewed representation of hospitalized patients in HGI datasets. Indeed, non-hospitalized-specific SNPs did not reach genome-wide significance, likely reflecting this bias; thus a relative weaker association threshold was used to prioritize mild-COVID specific SNPs for future investigation of long COVID. Second, the TWAS conducted using S-MetaXcan are constrained by the limited tissue specificity of available GTEx datasets and COVID related GWASs. Despite prioritizing 7 loci, comprising *HLA-DQA1*, *HLA-A*, *HLA-C*, *DPP9*, *CCR1*, *THBS3*, *GSDMA/B*, that emerged as promising TWAS hits for long COVID, the potential absence of COVID-19-specific tissues, such as nasopharyngeal or immune-relevant sites, may hinder the detection of tissue-specific effects pertinent to acute-phase COVID-19 or long COVID. Additionally, while our approach offers new insights into long COVID, we acknowledge that proxy-based inference carries inherent uncertainty. The overlaps observed with long COVID loci are suggestive rather than confirmatory or casual. Nonetheless, the concordance of TWAS-implicated genes and pheno-TWAS-trait enrichment supports the biological plausibility association between acute-phase COVID-19 and long COVID. Another limitation is that we leveraged acute COVID-19 GWAS data to prioritize SNPs and its adjacent genes for future investigation of long COVID; these results are only potential resources without confirmation of their causal relationships with long COVID, which require further in-depth investigation in independent long COVID cohort or confirmed by other experimental validations. Also, it is necessary to pointed out that only *FOXP4* emerged as a genome-wide significant locus associated with long COVID based on integrative analyses of COVID-19/long COVID GWAS/TWAS; the differential definitions of long COVID cases and controls coupled with the heterogeneity of long COVID may be the underlying reason for the low power of long COVID GWASs. Also, many prioritized loci demonstrated substantially weaker association signals in long COVID GWASs at both the SNP and imputed gene-expression levels compared with acute-phase COVID-19 GWASs; consequently, extensive downstream functional interpretation, including pathway enrichment and regulatory network analyses, may be premature at the current stage because such analyses could over-interpret loci lacking strong replication in long COVID cohorts. Future studies incorporating larger long COVID cohorts, refined clinical phenotyping, and additional functional genomics datasets, including cell-type-specific eQTL and epigenomic resources, will be important to validate the biological relevance of these candidate loci and refine causal SNP-to-gene relationships. Finally, our prioritized SNPs/loci would be helpful for researchers interested on long COVID, however, further colocalization or fine-mapping is strongly suggested to uncover or confirm the causal genetic factors to long COVID in the future.

## Data Availability

All data produced in the present study are available upon reasonable request to the authors.

## ACKNOWLEDGMENTS

We are thankful to all researchers who participated and contributed to the HGI and GRASP project, as their generosities in freely sharing these COVID-19 hospitalization GWASs and other COVID-19/long COVID related GWASs are essential for the current investigation.

## DECLARATION OF INTERESTS

The authors declare no competing interests.

## AUTHOR CONTRIBUTIONS

Z-S C conceived, analyzed the data and drafted the manuscript.

## FIGURE TITLES AND LEGENDS

**Figure 1.** Workflow for prioritizing genetic single nucleotide polymorphisms (SNPs) associated with either COVID-19 hospitalization or non-hospitalization or both as well as its association with sex-stratified COVID-19 hospitalization and long COVID An integrative analysis workflow is developed to analyze acute-phase COVID-19 GWAS, including HGI-C2 (SARS-CoV-2 infected cases vs. general population), HGI-B1 (hospitalized COVID-19 vs. non-hospitalized COVID-19), and HGI-B2 (hospitalized COVID-19 vs. general population), as well as the differential GWAS (HGI-B1-vs-B2) generated in current study by comparing effect sizes of each SNPs between HGI-B1 and HGI-B2. Three groups of top candidate SNPs, comprising “Severe COVID-19-Specific SNPs,” “SNPs Associated with Both Severe and Mild COVID-19,” and “Mild COVID-19-Specific SNPs,” are obtained by applying different filters of association p-values as demonstrated in the right part of the figure (read details in Materials and Methods section). These filters result in 62 candidate SNPs that are subsequently subjected to test associations with long COVID or sex-biased association with COVID-19 hospitalization. Further transcriptome-wide association studies and phenotype-wide transcriptome-wide association studies are conducted to investigate genes adjacent to these candidate SNPs and their association with acute-phase COVID-19 and long COVID.

**Figure 2.** Candidate SNPs associated with COVID-19 hospitalization or both hospitalization and non-hospitalization (A-B) 42 top prioritized candidate SNPs, with 27 SNPs specifically associated with hospitalization risk (left panel) and 15 showing common risk patterns across both hospitalized and non-hospitalized cases of COVID-19 (right panel). These SNPs were selected based on the analysis workflow illustrated in Fig. 1, and each SNP is centered within a 1Mb window in a local Manhattan plot to highlight the genomic context for its association with acute-phase COVID-19. Key genes near these SNPs, including *FOXP4*, a locus previously implicated as a risk locus for both acute-phase COVID-19 and long COVID, and other notable COVID-19-associated genes (*ACE2*, *OAS3*, *JAK1*, *IFNA2*, *DDP9*, and *ABO*), are annotated to emphasize their potential relevance to COVID-19. (C) Forest plot of 42 SNPs identified from COVID-19 GWAS on susceptibility (HGI-C2) and hospitalization (HGI-B1 and HGI-B2), showing suggestive (*P <* 1×10^-7^) or genome-wide significant associations (*P <* 5×10^-8^). SNPs are grouped as follows: (1) hospitalization-specific, genome-wide significant (Hosp-specific, *P <* 5×10^-8^); (2) common across cases, genome-wide significant (Common, *P <* 5×10^-8^); (3) hospitalization-specific, suggestive or non-significant (hosp-specific, *P* = [5×10^-8^, 1]); (4) common across cases, suggestive or non-significant (Common, *P* = [5×10^-8^, 1]). Colored dots represent odds ratios (ORs) with 95% confidence intervals (CIs) as error bars. ORs with CIs overlapping the vertical line (OR = 1) indicate no significance.

**Figure 3.** Evaluation of top genes adjacent to prioritized candidate SNPs (n=62) in TWAS of COVID-19 susceptibility, hospitalization, and long COVID (A) Bar plot showing the top genes adjacent to prioritized candidate SNPs based on TWAS for COVID-19 susceptibility and hospitalization, as well as long COVID. Genes are categorized by the GTEx tissue in which they display the most significant association. Besides rs17078348, other two SNPs have multiple genes with the smallest TWAS *P* are highlighted. The reference line indicates the arbitrary prioritization *P* = 3.5×10^-7^ (see Material and Methods for the derivation of this multiple testing adjustment). (B) Heatmap depicting the significance of gene-tissue associations for prioritized SNP-adjacent genes across 48 GTEx tissues. Each cell represents the *P* for a specific gene in a given tissue, with a color gradient indicating statistical significance: gray (*P* > 0.05), yellow (*P <* 0.05), and red (*P <* 1×10^-6^); for the sake of clear visualization in the heatmap, all associations with *P* <1×10^-6^ were truncated to be *P =* 1×10^-6^. Genes are sorted by TWAS names, and GTEx tissues are clustered to highlight patterns of tissue specificity.

## SUPPLEMENTAL INFORMATION

**Document S1. Figures S1–S24**

**Table S1. Association signals for these 62 prioritized candidate SNPs in Long COVID GWASs from HGI**

**Table S2. Association signals for SNPs adjacent to these 62 prioritized candidate SNPs in Long COVID GWASs from HGI**

**Table S3. Association signals for published long COVID SNPs across 4 HGI COVID-19 GWASs**

**Table S4 Top genes associated with acute-phase COVID-19 susceptibility and hospitalization across 48 GTEx tissues in transcriptome-wide association studies (TWAS)**

**Table S5. Long COVID association signals for the adjacent genes of these 62 candidate SNPs that passed the nominal significance in the transcriptome-wide association studies (TWAS) of COVID-19 hospitalization and susceptibility**

**Table S6. Adjacent genes of these 62 candidate SNPs that passed the nominal significance threshold in the phenome-wide transcriptome association studies (Pheno-TWASs)**

**Table S7. Transcriptome-wide association studies (TWAS) curated and used in current study**

## Notes

### Competing Interest Statement

The authors have declared no competing interest.

### Funding Statement

This study did not receive any funding.

### Author Declarations

All COVID-19 GWAS data used in the manuscript are freely available at https://www.covid19hg.org/.

### Summary of Updates

Revise some minor errors in methods and figure legends.

