## Supplementary document for "Integrative Genome-Wide Association Studies of COVID-19 Susceptibility and Hospitalization Reveal Risk Loci for Long COVID" (from another version of this preprint: version 5, 2026-05-13)

### SUPPLEMENTAL FIGURES

**Figure S1. QQ plots for four COVID-19 GWASs from HGI**

**Figure S2. Local Manhattan plots for candidate SNPs more likely associated with hospitalized than non-hospitalized COVID-19**

**Figure S3. Local Manhattan plots for candidate SNPs likely associated with both hospitalized and non-hospitalized COVID-19**

**Figure S4. Evaluation of prioritized candidate SNPs that tend to be either more likely associated with COVID-19 hospitalization or commonly associated with both hospitalization and non-hospitalization of COVID-19 in sex-biased COVID-19 hospitalization GWAS from UK Biobank**

(A-B) Manhattan plots and QQ plots for sex-stratified COVID-19 hospitalization GWASs, as well as the differential effect size GWAS between females and males. (C) Local Manhattan plot for these 42 SNPs separated into two groups, including candidate SNPs that tend to be specific in hospitalized cases and that are more frequent in both hospitalized and non-hospitalized cases. All association signals shown in the figure are within a 100Mb genomic window for each candidate SNP residing in the middle. Genes close to these 42 SNPs are added into the upper track. (C) Forest plots for these 42 candidate SNPs in sex-stratified COVID-19 hospitalization GWASs, as well as the differential effect size GWAS between females and males. Odd Ratios (ORs) and the corresponding 95% confidence intervals (CI) of these SNPs are illustrated, with SNPs showing nominal significant associations among female or male COVID-19 patients are colored in dark red and indicated by ‘*’. Note that there are 8 and 6 SNPs showing sex-biased associations with females and males, respectively, meanwhile, only one SNP rs17078348 mapped to *SLC6A20* displaying nominally significant association signals in both sexes. The genomic position of rs190509934 is used to collect all other SNPs around it, as it is not included in the two UKB sex-stratified COVID-19 hospitalization GWASs.

**Figure S5. Local Manhattan plots for candidate SNPs either more likely associated with COVID-19 hospitalization or commonly associated with both hospitalization and non-hospitalization of COVID-19 in sex-biased COVID-19 hospitalization GWAS from UK Biobank**

**Figure S6. Local Manhattan plots for candidate SNPs specifically associated with non-hospitalized COVID-19**

**Figure S7. Evaluation of candidate SNPs associated with non-hospitalized COVID-19 in sex-biased COVID-19 hospitalization GWAS from UK Biobank**

(A) Local Manhattan plots for candidate SNPs identified exclusively in the non-hospitalized COVID-19 group in sex-stratified COVID-19 hospitalization GWAS (the differential effect size GWAS between females and males put on the top, female GWAS added in the middle, and male GWAS shown at the bottom). (B) Forest plots displaying the odds ratios (ORs) and its corresponding 95% confidence intervals (CIs) of these SNPs in non-hospitalized COVID-19 patients from the sex-stratified GWAS (male results on the top and female results at the bottom). SNPs with nominally significant associations in either female or male groups are highlighted in dark red and marked with an asterisk (*). Notably, three SNPs exhibit male-biased associations with COVID-19 hospitalization. (C-H) Detailed local Manhattan plots for six prioritized SNPs specifically identified in the non-hospitalized COVID-19 group within the sex-stratified GWAS.

**Figure S8. Local Manhattan plots for SNPs in the SNP category “Severe COVID-19-Specific SNPs” across 4 long COVID GWASs**

Each target SNP is located at the center of each local Manhattan plot, and its linked gene is put along SNP rsid at the bottom of the figure.

**Figure S9. Local Manhattan plots for SNPs in the two SNP categories, including “Mild COVID-19-Specific SNPs” and “SNPs Associated with Both Severe and Mild COVID-19” across 4 long COVID GWASs**

Figure annotation is similar to that of Figure S8.

**Figure S10. Forest plots for 62 candidate SNPs and its adjacent SNPs passed the association threshold of *P*<0.001 in at least one of four long COVID GWASs**

SNPs adjacent to candidate SNPs that are prioritized from 4 HGI COVID-19 GWASs (colored according to its membership to 3 SNP categories in Figure 1) are indicated by adding “+” after the corresponding gene. “*” represents the SNP’s association with long COVID is nominally significant. To the sake of clarity, the OR is restricted to be less than 2. SNPs that are not include in specific long COVID GWASs show no OR and 95% CI in the forest plots.

**Figure S11. Evaluation of other published long COVID SNPs in the current HGI COVID-19 GWASs**

(A) Forest plots for 15 published long COVID SNPs in currently acute-phase COVID-19 GWASs (from left to right: HGI-B1, HGI-B2, and HGI-C2), with genes closed to each of them listed on the right-y-axis. Note: beside 3 long COVID risk SNPs reported by Chaudhary et al., other 12 long COVID SNPs were obtained by screening 83 previously reported long COVID SNPs by Taylor et al. (see Table S3) that show nominally significant association in at least one of the 3 acute-phase COVID-19 GWASs. (B-G) Local Manhattan plots illustrate published long COVID SNPs approximate to these top candidate SNPs identified in current study with each candidate SNP is located in the center of each genomic window; GWAS labels from bottom to top in each panel are HGI-B1, HGI-B2, HGI-B1-vs-B2, and HGI-C2, respectively.

**Figure S12. Total number of genes adjacent to prioritized candidate SNPs (n=62) as well as total number of genes that were tested and passed different association thresholds in COVID-19 transcriptome-wide association studies (TWASs)**

Bar plots illustrating the number of genes adjacent to prioritized candidate SNPs (n=62) as well as number of adjacent genes tested and passed different association *P* thresholds in TWASs. Note that the 62 SNPs are colored according to its corresponding SNP category as depicted in Figure 1. There are 4 acute-phase COVID-19 related TWASs and 4 long COVID TWASs performed across 48 GTEx tissues.

**Figure S13. Transcriptome-wide association studies (TWAS) for COVID-19 susceptibility, hospitalization, and long COVID**

(A) Top genome-wide significant TWAS-identified genes associated with COVID-19 susceptibility and hospitalization, annotated by their best significant tissue expression association (based on 48 GTEx tissues). No genes reached genome-wide significance in TWAS analyses of long COVID-related GWASs. (B) Manhattan plots depicting TWAS results for COVID-19 susceptibility, and hospitalization (HGI datasets), as well as long COVID GWASs (LongCOVID-N1, LongCOVID-N2, LongCOVID-W1, and LongCOVID-W2) derived from four distinct case-control designs. Each dot represents a gene, with its genomic position along the x-axis and -log10(*P*) on the y-axis.

**Figure S14. Evaluation of top genes emerged from transcriptome-wide association studies (TWAS) of COVID-19 susceptibility and hospitalization in the corresponding long COVID-related TWASs**

Heatmap displaying the significance (*P*) of top genes identified in TWAS analyses of COVID-19 susceptibility and hospitalization across multiple tissues in long COVID-related TWASs. Each cell represents the *P* for the association of a gene with long COVID in a specific tissue from GTEx database. The color gradient denotes statistical significance: blue (*P*>0.05), yellow (*P*<0.05), and red (*P*<1x10^-6^). Rows are sorted by genes, and columns represent tissues, allowing visualization of tissue-specific patterns of gene expression relevance.

**Figure S15. Other top genes adjacent to prioritized candidate SNPs (n=29) in long COVID-19 transcriptome-wide association studies (TWASs)**

Bar plots illustrating the best *P* for genes adjacent to prioritized candidate SNPs (n=29) in 4 long COVID TWASs across 48 GTEx tissues, which is a supplement to Figure 4.

**Figure S16. Evaluation of top genes adjacent to prioritized candidate SNPs (n=62) in transcriptome-wide association studies (TWASs) of long COVID across 48 GTEx tissues**

(A) Bar plot showing the number of SNP-gene pairs passed the nominal TWAS association threshold in GTEx tissues. Note that SNP-gene pairs are defined by combining prioritized candidate SNPs with its adjacent genes. (B) Heatmap depicting the significance (*P*) of SNP-gene pairs across 48 GTEx tissues in long COVID TWASs. Each cell represents the *P* for a specific gene in a given tissue, with a color gradient indicating statistical significance: white (*P*>0.05), yellow (*P*<0.05), and red (*P*<1E-6). Genes are sorted by SNP-gene pairs and TWAS names.

**Figure S17. Evaluation of genes adjacent to prioritized candidate SNP rs17219281 (*HLA-DQA1*) in phenome-transcriptome-wide association studies (Pheno-TWAS) across 48 GTEx tissues**

To the sake of clarity, heatmap only depicting the significance of SNP-gene pairs with *P*<1x10^-60^ across 48 GTEx tissues in TWASs. Each cell represents the *P* for a specific gene in a given tissue, with blank cell indicating the *P* not passed the threshold of *P*<1x10^-60^.

**Figure S18. Evaluation of genes adjacent to prioritized candidate SNP rs9260038 (*HLA-A*) in phenome-transcriptome-wide association studies (Pheno-TWAS) across 48 GTEx tissues**

Annotation is similar to heatmap as that in Figure S17.

**Figure S19. Evaluation of genes adjacent to prioritized candidate SNP rs1634761 (*HLA-C*) in phenome-transcriptome-wide association studies (Pheno-TWAS) across 48 GTEx tissues**

Annotation is similar to heatmap as that in Figure S17.

**Figure S20. Evaluation of genes adjacent to prioritized candidate SNP rs2834164 (*IFNAR2*) in phenome-transcriptome-wide association studies (Pheno-TWAS) across 48 GTEx tissues**

To the sake of clarity, the color gradient of heatmap denotes statistical significance: blue (*P*>0.05), yellow (*P*<0.05), and red (*P*<11x10^-6^). Rows are sorted by genes, and columns represent TWAS names, allowing visualization of tissue-specific patterns of gene expression relevance across 48 GTEx tissues in TWASs.

**Figure S21. Evaluation of genes adjacent to prioritized candidate SNP rs9916158 (*GSDMA/B*) in phenome-transcriptome-wide association studies (Pheno-TWAS) across 48 GTEx tissues**

Heatmap legend as that in Figure S20

**Figure S22. Evaluation of genes adjacent to prioritized candidate SNP rs12602210 (*BPTF*) in phenome-transcriptome-wide association studies (Pheno-TWAS) across 48 GTEx tissues**

Heatmap legend as that in Figure S20

**Figure S23. Evaluation of genes adjacent to prioritized candidate SNP rs79611697 (*CCDC171*) in phenome-transcriptome-wide association studies (Pheno-TWAS) across 48 GTEx tissues**

Heatmap legend as that in Figure S20

**Figure S24. Evaluation of genes adjacent to prioritized candidate SNP rs12660421 (*FOXP4*) in phenome-transcriptome-wide association studies (Pheno-TWAS) across 48 GTEx tissues**

Heatmap legend as that in Figure S20
